# Network and receptor architectures shape brain morphometry in addiction

**DOI:** 10.64898/2026.04.03.26348150

**Authors:** Foivos Georgiadis, Beatrice A. Milano, Sara Larivière, Kent E. Hutchinson, Vince Calhoun, Chiang-Shan Ray Li, Reza Momenan, Rajita Sinha, Dick Veltman, Ruth van Holst, Anneke Goudriaan, Maartje Luijten, Martine Groefsema, Henrik Walter, Tristram Lett, Reinout Wiers, Lianne Schmaal, Julianne Flanagan, Bernice Porjesz, Jonathan Ipser, Justin Boehmer, Nicola Canessa, Ramiro Salas, Edythe London, Martin Paulus, Dan Stein, Samantha Brooks, Liesbeth Reneman, Anouk Schrantee, Francesca Filbey, Rob Hester, Murat Yücel, Valentina Lorenzetti, Nadia Solowij, Rocío Martín-Santos, Albert Batalla, Janna Cousijn, Edith Pomarol-Clotet, Eduardo A. Garza-Villarreal, Marco Leyton, Elliot Stein, Cleo L. Crunelle, Anne M. Kaag, Antonio Verdejo-Garcia, John J. Foxe, Kathleen T. Brady, Aimee McRae-Clark, Alain Dagher, Amelie Haugg, Marc Walter, André Schmidt, Anne Lingford-Hughes, Louise M. Paterson, Angelica M. Morales, Dara G. Ghahremani, Chuan Fan, Etna J. E. Engeli, Marcus Herdener, Boris B. Quednow, Erich Seifritz, Philipp Homan, Marco De Pieri, Silke Bachmann, Daniele Zullino, Justine Y. Hansen, Bratislav Misic, Sophia I. Thomopoulos, Paul M. Thompson, Devarshi Pancholi, Anthony Juliano, Hugh Garavan, Sofie L. Valk, Boris C. Bernhardt, Matthias Kirschner, ENIGMA Addiction Working Group

## Abstract

Substance use disorders (SUD) are associated with widespread brain structural alterations, but the principles governing their regional distribution remain unclear. We tested whether SUD-related morphometric differences are shaped by normative connectivity and neurotransmitter receptor architecture. We analysed T1-weighted MRI data from 2,782 individuals with SUD (mean age 34.6 years [SD 11.2]; 33.3% female) and 1,951 healthy controls (mean age 33.8 years [SD 12.0]; 37.5% female) across 51 ENIGMA Addiction Working Group sites, spanning alcohol, amphetamine, cannabis, cocaine, nicotine and opioid use disorders. Cortical thickness (68 regions) and subcortical volume (14 structures) were quantified using harmonized ENIGMA protocols. Case-control maps were related to normative functional and structural connectomes through hub-vulnerability and epicenter analyses, and to 20 PET-derived receptor-density maps using partial least squares analysis.

Across substances, SUD was associated with lower cortical thickness and subcortical volume involving frontal, parietal, temporal and limbic regions. Alterations overlapped with cortical hubs and converged on cortical and subcortical epicenters, with findings robust across six substances, biological sex and analytical approaches. SUD epicenters showed strong spatial overlap with those of schizophrenia and bipolar disorder, particularly in frontotemporal, parietal and striatal regions, distinct from mood, neurodevelopmental and obsessive-compulsive conditions. Cortical alteration patterns aligned with two neurochemical axes: one enriched in cannabinoid, opioid, metabotropic glutamate and histamine receptors, and another associated with lower dopaminergic and cholinergic marker density.

These findings indicate that addiction-related brain differences are organized by normative connectome and neurotransmitter architecture, providing a systems-level framework for identifying receptor- and circuit-based targets for intervention in addiction.

## Introduction

Substance use disorders (SUD) are a major and growing global health concern, affecting millions of individuals worldwide, contributing substantially to the overall burden of mental illness.^1,2^

SUD are understood to arise from a multifactorial interplay between environmental factors and biological vulnerability, including genetic risk.^3,4^ Converging evidence from neuroimaging, neuropathological and molecular studies has identified consistent differences in brain structure and function associated with SUD.^5–7^ Large-scale MRI case-control studies from the Enhancing NeuroImaging Genetics through Meta-Analysis (ENIGMA) Addiction Working Group have demonstrated robust cortical and subcortical morphometric differences, across multiple substances, including alcohol, cocaine, cannabis, nicotine and amphetamines.^8,9^ These alterations involve brain systems implicated in reward processing, salience attribution and learning.^10,11^

Structural differences have been linked to both the stage and the severity of addiction, with converging evidence indicating a dose-response relationship between cumulative substance exposure and macroscopic neuroanatomical differences; for example, progressively lower cortical thickness, smaller subcortical volumes, reduced white-matter integrity, and advanced brain-age, with the strongest evidence in alcohol and cocaine use,^12–17^ extending to amphetamines, cannabis, nicotine and opioids.^18–20^ Mechanistically, these patterns plausibly reflect combined neurotoxic and neuroimmune processes such as neuronal injury, white matter compromise, microvascular damage, and neuroinflammation,^21–23^ moderated by genetic liability and early-life adversity.^6,24^

Despite this growing evidence, the principles that organize the spatial distribution of SUD-related morphometric differences remain poorly understood. Prior work has shown that structural abnormalities in brain disorders do not arise randomly but are spatially coordinated, reflecting the brain’s intrinsic organizational architecture.^25–29^ This raises the question of whether network-based models of brain connectivity can explain why some regions are more affected than others.

Two complementary models from network neuroscience are particularly relevant. The hub vulnerability hypothesis posits that hub regions, i.e., highly interconnected brain regions that are central for information integration and characterized by high metabolic costs, are disproportionately susceptible to damage.^30–33^ Evidence for this model first emerged in neurodegenerative disorders, where hub regions such as the posterior cingulate cortex and precuneus were shown to be preferentially affected in Alzheimer’s disease.^34^ Subsequent studies extended this principle to psychiatric disorders, including schizophrenia.^26^ In addition to cortical hubs, current circuit models of addiction emphasize that dysregulated communication between limbic/striatal hubs and prefrontal control regions underlies compulsive drug seeking and impaired self-regulation, underscoring the relevance of subcortico-cortical connectivity.^35,36^ The epicenter model posits that pathological processes can be traced back to specific regions acting as epicenters; epicenters are regions whose normative connectivity profile matches the observed spatial distribution of structural differences, with spread proposed to occur along connected pathways.^28,33,37^ Initially developed in neurodegenerative disorders^38^ such as Parkinson’s^39^ and Alzheimer’s disease, and frontotemporal dementia,^40^ this framework has since been extended to epilepsy and psychiatric disorders.^25–27,41–43^

A distinct feature of SUD compared to other psychiatric disorders is that neurobiology interacts with the effects of substances.^6^ Psychoactive substances act on distinct neurotransmitter receptor systems, whose cortical distribution is closely aligned with the brain’s network topology.^44,45^ These receptor landscapes could plausibly act as molecular scaffolds that constrain the spread of pathology along functional circuits. Yet the principles organizing the spatial distribution of morphometric differences in addiction, and the extent to which network connectivity and neurochemical architecture jointly constrain these patterns, remain to be established.

Here, we integrate these network and molecular perspectives to test the hypothesis that structural brain differences in SUD are shaped by normative connectivity and neurotransmitter receptor distributions. Using structural brain MRI data from 2,782 individuals with SUD and 1,951 healthy controls from the ENIGMA Addiction Working Group, we (i) characterized morphometric differences across individuals with SUD and within six substance use categories, (ii) tested whether these differences preferentially affect hubs as well as whether they follow the connectivity profiles of specific epicenter regions, (iii) examined cross-disorder similarity with other major psychiatric conditions, and (iv) linked SUD-related morphometric difference patterns to 20 PET-based neurotransmitter receptor and transporter maps (e.g., dopamine, serotonin, GABA, acetylcholine, opioid and histamine systems). We further performed complementary robustness and individual-level analyses to confirm stability and generalizability of our findings.

## Materials and methods

The overall analytic workflow, including data integration, morphometric modelling, and network-based analyses, is summarized in **Fig. 1**. All network-level modelling was conducted with Python 3.10.13 using the open-source ENIGMA Toolbox.^46^ Additional statistical analyses were carried out in R 4.3.0.

**Fig. 1.**
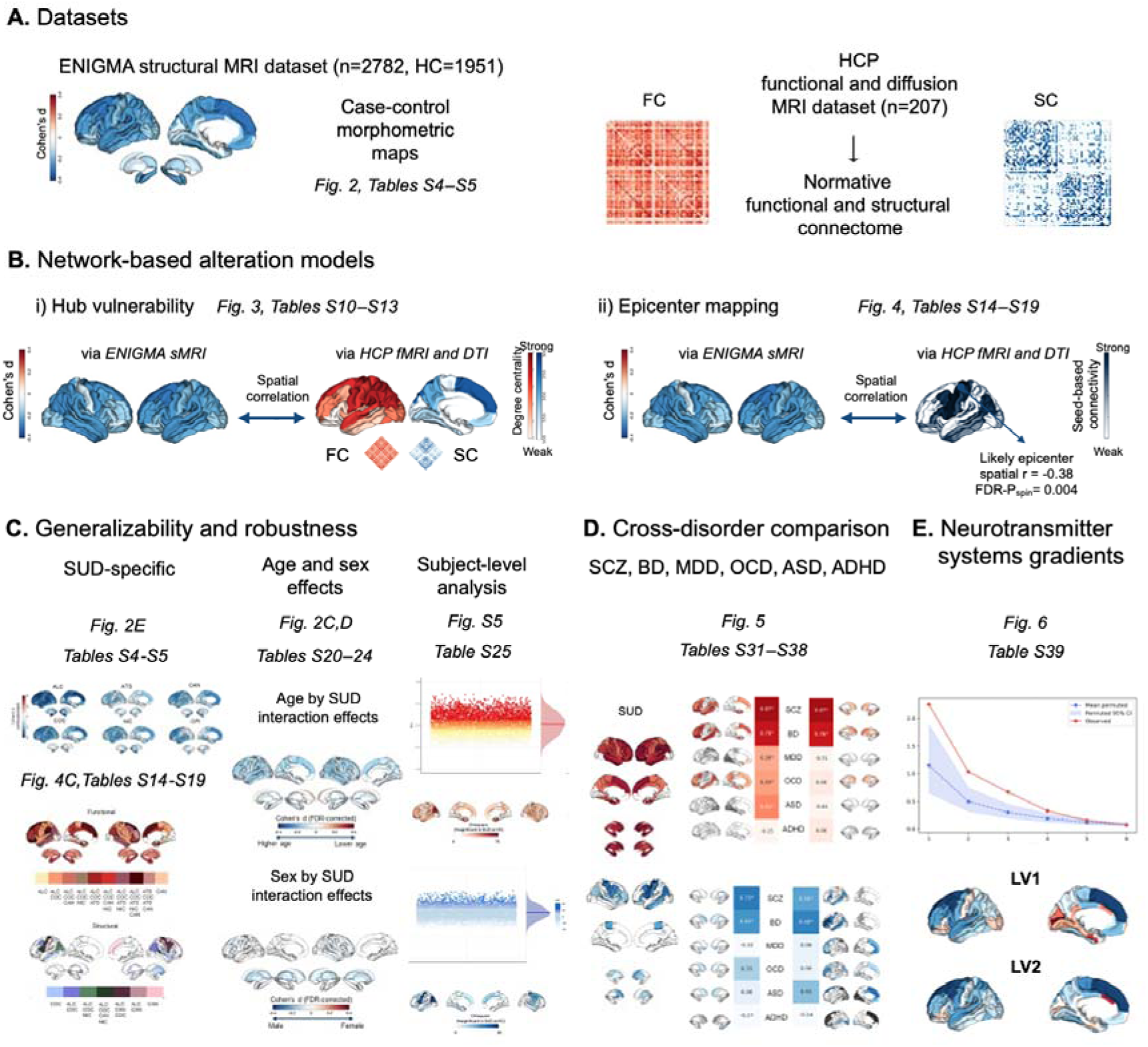
Overview of the datasets and analysis pipeline. (**A**) ENIGMA Addiction Working Group structural MRI dataset (*n* = 2,782 individuals with SUD and *n* = 1,951 healthy controls), including six substance categories: alcohol, amphetamines, cocaine, cannabis, nicotine and opioids. Case–control cortical thickness and subcortical volume alteration maps were generated for each substance category and for the combined SUD group. (**B)** Normative functional and structural connectomes from the Human Connectome Project (*n* = 207) were used to test two network-based models of SUD-related brain alterations: hub vulnerability and epicenter mapping. (**C**) Robustness and generalizability analyses, including substance-specific profiles, SUD-by-age, SUD-by-sex interaction effects, and subject-level analyses. (**D**) Cross-disorder comparisons of SUD-related maps with those from schizophrenia, bipolar disorder, major depressive disorder, obsessive-compulsive disorder, autism spectrum disorder, and attention-deficit/hyperactivity disorder. (**E**) Association of neurotransmitter density with cortical alterations and the connectome organization in SUD. Abbreviations: ADHD = attention-deficit/hyperactivity disorder; ASD = autism spectrum disorder; BD = bipolar disorder; DTI = diffusion tensor imaging; ENIGMA = Enhancing NeuroImaging Genetics through Meta-Analysis; FC = functional connectivity; FDR = false discovery rate; fMRI = functional MRI; HC = healthy controls; HCP = Human Connectome Project; LV = latent variable; MDD = major depressive disorder; MRI = magnetic resonance imaging; n = number of participants; OCD = obsessive-compulsive disorder; SC = structural connectivity; SCZ = schizophrenia; sMRI = structural MRI; SUD = substance use disorder.

### ENIGMA Participants

We analysed structural neuroimaging data from 2,782 individuals with SUD (mean age ± standard deviation [SD] = 34.6 ± 11.2 years; range: 18–67; 33.3% female, *n =* 926) and 1,951 healthy controls (mean age = 33.8 ± 12.0 years; range: 18–75; 37.5% female, *n =* 731) collected across 51 international sites from the ENIGMA Addiction Working Group (Supplementary **Table 1**; Supplementary Fig. **1**). Participants with SUD met diagnostic criteria for one or more of six primary categories: alcohol, amphetamines, cannabis, cocaine, nicotine and opioids. A total of 2,312 individuals (mean age = 34.7 ± 11.3 years; range: 18–67; 33.9% female, *n =* 783) met criteria for a single SUD diagnosis without comorbidity of another SUD and were included in substance-specific analyses (alcohol: *n* = 902; amphetamines: *n* = 178; cannabis: *n* = 286; cocaine: *n* = 278; nicotine: *n* = 600; opioids: *n* = 68). Diagnoses were confirmed at each site using Diagnostic and Statistical Manual of Mental Disorders, Fourth Edition (DSM-IV) or International Classification of Diseases, 10th Revision (ICD-10), criteria. Site-level demographic details are reported in Supplementary **Table 2**. Individuals were excluded if they had a lifetime history of neurological disease, a current DSM-IV/DSM-5 axis I diagnosis other than depressive and anxiety disorders, or any contraindication for MRI. Control subjects may have used addictive substances recreationally but were never diagnosed with any SUD. All studies were approved by local institutional review boards, and all participants provided written informed consent.

Primary analyses focused on the combined SUD (n = 2,782) and control sample (n = 1,951), including all six substance categories, in line with prior ENIGMA work.^8^ We then conducted substance-specific analyses, excluding individuals with more than one SUD, as secondary analyses to test the consistency of main findings across the individual SUD categories.

#### Image acquisition and processing

High-resolution three-dimensional volumetric T1-weighted structural brain MRI scans were acquired at each site using local acquisition protocols (Supplementary **Table 3**). Imaging data were processed using the ENIGMA standard structural MRI pipeline with FreeSurfer version 5.3.^47,48^ Cortical thickness was extracted from 68 bilateral regions using the Desikan-Killiany atlas.^49^ Volumetric measures were obtained for 14 subcortical grey matter regions, including bilateral amygdala, thalamus, caudate, putamen, pallidum, nucleus accumbens and bilateral hippocampi. All sites adhered to ENIGMA’s standardized quality control procedures (http://enigma.usc.edu/protocols/imaging-protocols).

#### Statistical mega-analysis

To assess morphometric differences in cortical thickness (CT) and subcortical volume (SV) between individuals with SUD and healthy controls, we applied mega-analytical linear models across all 68 cortical and 14 subcortical regions. Morphometric data were harmonized across the 51 sites using ComBat, an empirical Bayes-based method for correcting batch effects.^50,51^ After harmonization, linear models were fit using the lm function in R to compare the average morphometric difference profiles for CT and SV in the combined sample of patients with SUD (*n =* 2,782) relative to controls (*n =* 1,951), correcting for age, sex, and intracranial volume (ICV) for both cortical and subcortical measures, similar to previous work,^8^ to ensure comparability. In our substance-specific analysis individuals with single SUD (alcohol: *n* = 902; amphetamines: *n* = 178; cannabis: *n* = 286; cocaine: *n* = 278; nicotine: *n* = 600; opioids: *n* = 68) were compared to controls (*n =* 1,951), applying the same linear model parameters. In a separate robustness analysis, an age-squared term was included as an additional covariate. Findings were corrected for multiple comparisons using the false discovery rate (FDR) procedure (Supplementary **Tables 4–5**).^52^ Leave-one-out analyses confirmed that combined-SUD morphometric patterns were not driven by any single substance category (Supplementary Methods and Results; Supplementary **Tables 6–9**).

Finally, we applied principal component analysis (PCA) to assess shared variance across substance-specific morphometric maps and high correspondence between the brain pattern derived from the first principal component (PC1) of the six single SUD and the combined SUD case-control maps (Supplementary Results; Supplementary **Fig. 2B**).

#### Functional and structural connectivity matrix generation

Normative functional and structural connectivity matrices were obtained from the ENIGMA Toolbox^46^ based on resting-state functional MRI (fMRI) and diffusion MRI data from 207 unrelated healthy young adults (83 males; mean age ± SD = 28.7 ± 3.7 years) from the Human Connectome Project (HCP).^53^ For details see Supplementary Methods.

#### Hub vulnerability model

To test the hub vulnerability hypothesis,^30–33^ we correlated the normative functional and structural centrality maps derived from cortico-cortical and subcortico-cortical connectivity with the regional cortical thickness and subcortical volume alterations, respectively. Centrality was defined as weighted degree centrality, reflecting the total strength of functional or structural connections of each region, weighted by their respective magnitude.^30^ Unthresholded centrality maps were used to avoid arbitrary cut-offs. To correct for spatial autocorrelation, significance was assessed using 10,000 surface-based spin permutations for cortical data.^46,54,55^ For subcortical regions, a similar procedure was applied, but instead of projecting subcortical labels onto spheres as in the spin test, the labels were randomly shuffled (Supplementary **Tables 10–11**).^56^ As a robustness check, we replicated the analyses using numerous centrality measures, including strength, eigenvector, betweenness, and closeness centrality, which capture complementary aspects of nodal importance in the network (Supplementary **Tables 12–13)**.

#### Mapping disease epicenters

Epicenters were identified by correlating each region’s normative connectivity profile (i.e., each row in the normative functional or structural connectivity matrix, represented as a vector) with the vector of SUD-related morphometric alterations.^25,28,46^ Spatial autocorrelation was addressed using spin permutation tests (10,000 repetitions) for cortical regions and label shuffling (10,000 repetitions) for subcortical regions (Supplementary **Tables 14–15**).^26,46,57^ Regions were considered epicenters if their correlation survived FDR correction (*P* < 0.05; *n =* 68 cortical, *n =* 14).

##### Substance-specific network modelling and cross-correlation structure

Substance-specific analyses were restricted to individuals with one primary SUD and compared with the pooled control sample to assess cortical thickness and subcortical volume differences across the six substance groups (Supplementary **Tables 4–5**), with robustness to comorbid SUD and non-pooled control samples assessed separately. Based on these substance-specific morphometric maps, we tested hub vulnerability, performed functional and structural epicenter mapping, assessed cross-substance spatial similarity, and identified epicenters across substances, correcting for spatial autocorrelation and multiple testing where appropriate (Supplementary **Tables 10–19**).

#### Robustness and generalizability analyses

We performed robustness and generalizability analyses including SUD-by-age and SUD-by-sex interactions, subject-level cortical abnormality models, age-harmonization bootstraps, sample-size equalization analyses, spatial stability analyses and alternative control-sampling strategies. Details are provided in the Supplementary Methods and Supplementary **Tables 20–30**.

#### Transdiagnostic disorder comparison

We used meta-analytical structural brain maps from the ENIGMA consortium for schizophrenia,^58,59^ bipolar disorder,^60,61^ major depressive disorder,^62,63^ obsessive-compulsive disorder,^64,65^ attention-deficit/hyperactivity disorder ^66,67^ and autism spectrum disorder.^68^ For each disorder, functional and structural epicenters were computed using the same mapping procedure as our SUD analysis (Supplementary **Tables 31–34**).

These were then compared with SUD epicenter maps to calculate cross-disorder spatial similarity (Supplementary **Tables 35–36**), with significance determined using spin tests and FDR correction. Only significant epicenters in both SUD and each of the disorders were included in the overlap analyses (Supplementary **Tables 37–38**).

#### Association of neurotransmitter density with cortical alterations and the connectome in SUD

We analysed 20 PET-derived cortical receptor maps aligned to the Desikan-Killiany atlas (68 regions; full list in Supplementary **Table 39)** derived from previously published PET datasets ^45^ and original receptor-specific PET studies. All receptor maps were z-scored across regions and assembled into a neurotransmitter matrix. SUD-related cortical alterations were quantified as regional Cohen’s *d* effect sizes for each of the six substance groups. These maps were also z-scored and organized into a second matrix representing SUD effects across the same 68 cortical regions. To identify shared spatial structure between neurotransmitter distributions and SUD-related cortical differences, we used partial least squares (PLS) analysis. This multivariate approach identifies pairs of latent variables (LVs) that maximize the covariance between the neurotransmitter and SUD matrices. The analysis yields (i) weights for each neurotransmitter receptors (*U*) and each SUD maps (*V*); (ii) region-wise LV scores, computed as the projection of each standardized matrix onto the corresponding weight vector; and (iii) loadings, computed as the Pearson correlation between each variable and the latent variable score of the same data block (i.e. individual receptor density maps with the receptor scores and single SUD cortical Cohen’s *d* maps with the SUD scores). Because the SUD-side weights were negative for all six substances, lower SUD-side scores correspond to more negative Cohen’s *d* values. To assess statistical significance, we performed 10,000 spin permutation tests in which the spatial order of the SUD matrix was permuted, preserving spatial autocorrelation, while the neurotransmitter matrix was kept intact. For each permutation, the full PLS solution was recomputed, generating a null distribution against which the observed latent variables were evaluated. We considered an LV significant if its associated singular value exceeded the 95% range of the spin permutation distribution. To evaluate the robustness of individual neurotransmitter and SUD loadings, we expressed each observed loading as a z-score relative to the spin permutation null distribution of loadings, with the sign of each permuted component aligned to the corresponding observed component prior to computing the null distribution. We correlated the SUD-side LV scores with degree centrality derived from normative functional and structural connectivity matrices from the ENIGMA Toolbox^46,54^ to examine whether the spatial expression of significant LVs is constrained by the functional and structural connectomes. Significance was assessed using spin permutation tests (10,000 repetitions).

## Results

### Structural differences across SUD

We compared structural neuroimaging data from 2,782 individuals with at least one SUD and 1,951 healthy controls across 51 international ENIGMA Addiction Working Group sites (**Fig. 2A**; Supplementary **Table 1**). Across the cortex, individuals with SUD showed widespread lower cortical thickness (**Fig. 2B**; Supplementary **Table 4**). After FDR correction, 65 of 68 cortical regions showed significant reductions (FDR-corrected *P* < 0.05), with the largest effects in frontal, parietal, and temporal regions, including the left superior frontal gyrus (Cohen’s *d* = −0.33), left caudal middle frontal gyrus (Cohen’s *d* = −0.31), and right inferior parietal cortex (FDR-corrected P < 0.05, Cohen’s *d* = −0.29). Subcortically, 11 of 14 structures showed lower values, with the strongest effect sizes in the hippocampus (−0.25/−0.24) and amygdala (−0.24/−0.21) (**Fig. 2B**; Supplementary **Table 5;** FDR-corrected *P* < 0.05). Effect size patterns were robust to alternative age specifications, with inclusion of a quadratic age term having negligible impact. SUD-related structural differences interacted with ageing, with more pronounced cortical, but not subcortical, case-control differences at older ages, consistent with age-dependent or cumulative effects (**Fig. 2C**; Supplementary **Tables 20–21**). Effects of biological sex were comparatively subtle and localized to subcortical regions (**Fig. 2D**; Supplementary **Tables 23–24**). To assess robustness of the combined-SUD phenotype and ensure results were not driven by any single substance, we conducted a leave-one-out analysis (Supplementary **Tables 6–7**) revealing consistent findings as reflected by high spatial correlations with the combined SUD map including all six single SUD (Supplementary **Tables 8–9**).

**Fig. 2.**
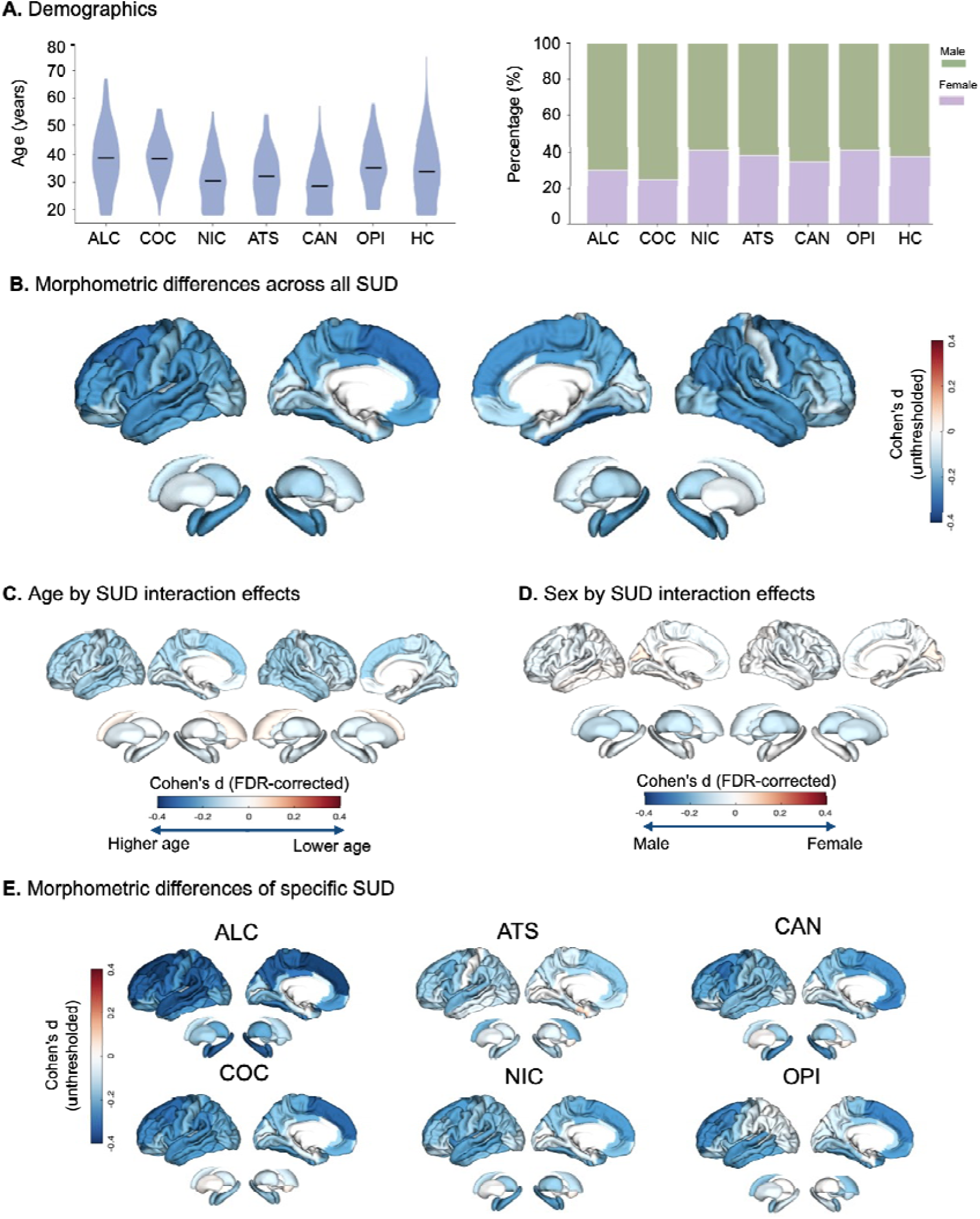
Morphometric differences in SUD. (**A**) Demographic characteristics of all groups. Age distributions are shown as mean ± SD (*n*) for controls and for individuals with a primary substance use disorder diagnosis (SUD): controls 33.8 ± 12.0 (*n =* 1,951), alcohol 38.8 ± 12.0 (*n =* 902), cocaine 38.5 ± 8.4 (*n =* 278), nicotine 30.4 ± 9.8 (*n =* 600), amphetamines 32.1 ± 10.1 (*n =* 178), cannabis 28.6 ± 9.2 (*n =* 286), and opioids 35.2 ± 9.8 (*n =* 68). Sex distributions (percentage female) were: controls 37.5%, alcohol 30.0%, cocaine 24.8%, nicotine 41.3%, amphetamines 38.2%, cannabis 34.6%, and opioids 41.2%. For more details, see Supplementary **Table 2**. (**B**) Unthresholded effect size maps of morphometric differences across the combined SUD phenotype, showing changes in cortical thickness and subcortical volume as quantified by Cohen’s *d* (see Supplementary **Tables 4–5**). (**C**) FDR-corrected maps of SUD-by-age interaction effects across the combined SUD phenotype, showing which cortical and subcortical regions are more severely affected in older versus younger individuals (see Supplementary **Tables 20–21**). (**D**) FDR-corrected maps of SUD-by-sex effects across all SUD categories, showing which cortical and subcortical regions are more severely affected in male versus female individuals (see Supplementary **Tables 23–24)**. (**E**) Unthresholded effect size maps of morphometric differences in specific SUD categories (alcohol, amphetamines, cannabis, cocaine, nicotine, opioids) for cortical thickness and subcortical volume, illustrating substance-specific variations in effect magnitude on a shared spatial topography (see Supplementary **Tables 4–5**). Abbreviations: ALC = alcohol use disorder; ATS = amphetamine-type stimulant use disorder; CAN = cannabis use disorder; COC = cocaine use disorder; FDR = false discovery rate; HC = healthy controls; NIC = nicotine use disorder; OPI = opioid use disorder; SUD = substance use disorder. Cohen’s d = standardized mean difference.

### SUD-specific structural differences

Substance-specific analyses revealed heterogeneous but substantially overlapping morphometric difference patterns (**Fig. 2E**; Supplementary **Tables 4–5**). Lower cortical thickness was most pronounced in alcohol use disorder (peak Cohen’s *d* = −0.47), followed by cocaine (−0.34), with nicotine (−0.28) showing intermediate effects that were less consistent across regions. Subcortical differences followed a similar ranking, with alcohol showing the most consistent volume reductions across nearly all subcortical structures (peak Cohen’s *d* = −0.34), followed by nicotine (−0.24). Cannabis use was associated with more circumscribed, limbic-predominant reductions, particularly in the hippocampus (−0.27) and amygdala. Amphetamine- and opioid-related effects did not survive FDR correction and were limited to small, non-significant trends (**Fig. 2E**; Supplementary **Tables 4–5**). Overall, substance-specific brain morphometric differences showed highly similar spatial patterns (Supplementary **Fig. 2A** and **B**).

### Functional and structural degree centrality predict regional susceptibility in SUD

According to the hub vulnerability hypothesis, brain regions with high topological centrality (hubs)^30^ are disproportionately susceptible to biological insults due to their central role in information integration, long-range communication, and high metabolic demands.^69^ We tested whether the magnitude of regional morphometric differences identified in our mega-analysis (**Fig. 2B**; Supplementary **Tables 4–5**) correlated with each region’s network centrality (**Fig. 3A**). Cortical regions exhibiting higher functional and structural centrality showed significantly lower cortical thickness in the combined SUD map (functional connectivity: *r* = –0.51, spin-permutation *P*-value (*P*_spin_) = 0.002; structural connectivity: *r* = –0.49, *P*_spin_ < 0.0001; **Fig. 3B**; Supplementary **Table 10**). In contrast, subcortical volume reductions were not significantly associated with subcortico-cortical degree centrality (both label-shuffling *P*-value (*P*_shuff_) ≥ 0.17; Supplementary **Table 11**).

**Fig. 3.**
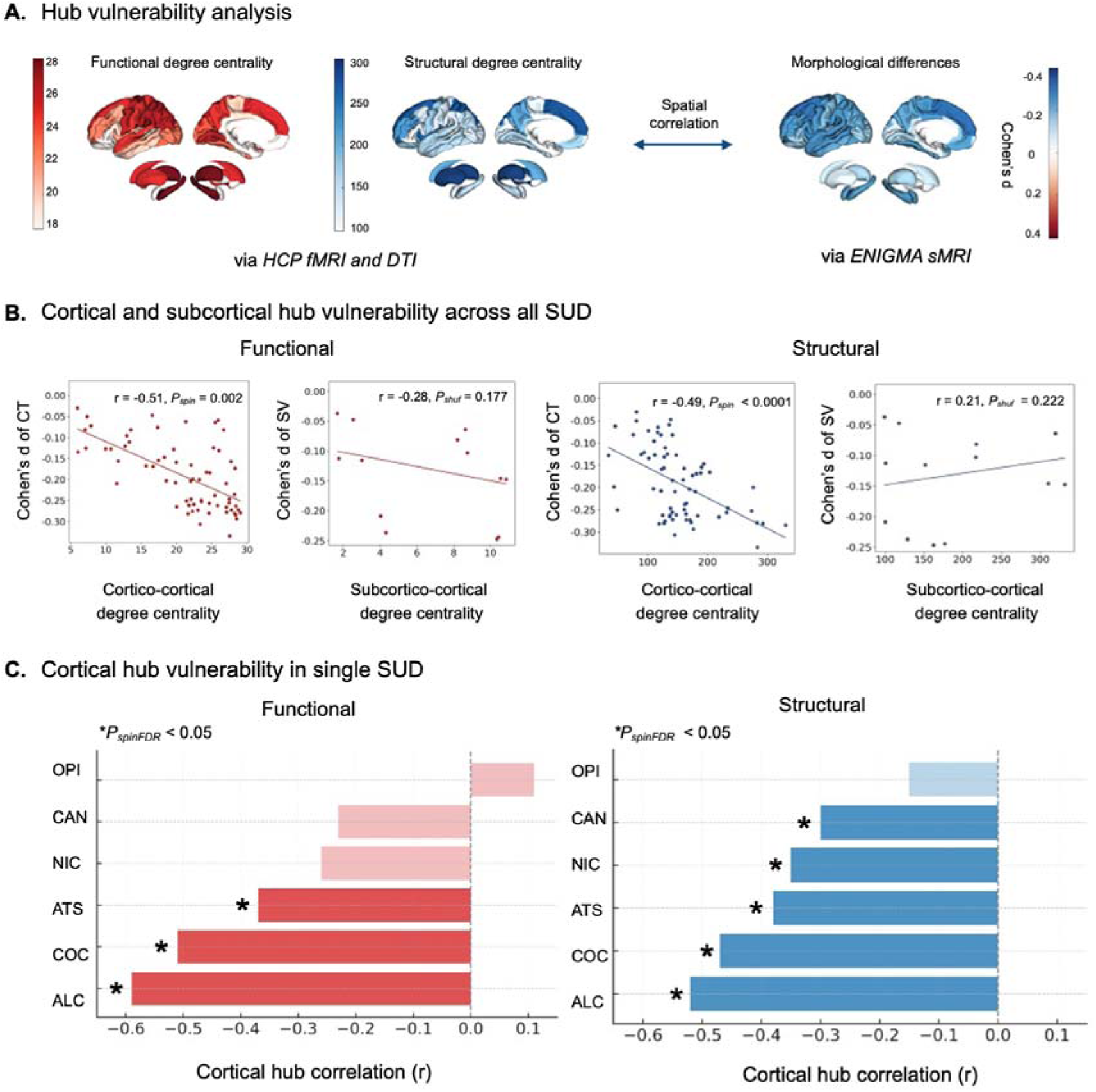
Hub vulnerability in functional and structural connectivity in SUD. (**A**) Left: HCP-derived normative functional and structural centrality, showing degree centrality for both functional and structural connectivity. Right: mega-analytic SUD morphometric map from our analysis. (**B**) Correlation plots show the spatial relationship between cortical thickness and subcortical volume differences (Cohen’s *d*) in the combined SUD phenotype and normative degree centrality, shown separately for functional vs structural and cortico-cortical vs subcortico-cortical connectivity. (**C**) For each single SUD, separate bar plots illustrate spatial correlations between cortical thickness differences (Cohen’s *d*) and HCP-derived functional (left, red) and structural (right, blue) cortico-cortical degree centrality. Cortical spatial correlations were assessed using spin-permutation tests (10,000 repetitions) to correct for spatial autocorrelation.^54^ Subcortical spatial correlations were assessed by randomly shuffling subcortical labels (10,000 repetitions). Significant correlations are shown in full colour and marked with an asterisk (FDR-P_spin_ < 0.05); faded bars denote non-significant correlations. Abbreviations: ALC = alcohol use disorder; ATS = amphetamine-type stimulant use disorder; CAN = cannabis use disorder; COC = cocaine use disorder; CT = cortical thickness; DTI = diffusion tensor imaging; ENIGMA = Enhancing NeuroImaging Genetics through Meta-Analysis; FDR = false discovery rate; HCP = Human Connectome Project; MRI = magnetic resonance imaging; NIC = nicotine use disorder; OPI = opioid use disorder; FDR-*P*_spin_ = spin-permutation *P*-value corrected for false discovery rate; P_shuff_ = label-shuffling P-value; *r* = Pearson correlation coefficient; sMRI = structural MRI; SUD = substance use disorder; SV = subcortical volume.

### Substance-specific hub vulnerability

We next examined whether the observed relationship between brain network centrality and morphometric differences in SUD varied across substance categories (**Fig. 2E**). In alcohol, cocaine and amphetamine use disorders, higher functional and structural degree centrality was associated with lower cortical thickness (all *r* < −0.30, FDR-*P*_spin_ < 0.05; **Fig. 3C**), whereas in nicotine and cannabis use disorders, higher structural centrality was associated with lower cortical thickness only (all *r*_struc_ ≤ −0.30, FDR-*P*_spin_ < 0.05; **Fig. 3C**; Supplementary **Tables 10–11**). Overall, cortical hub vulnerability was robust across SUD and consistent across centrality metrics (Supplementary **Tables 12–13**), whereas subcortico-cortical hub vulnerability was not significant.

### Disease epicenters of SUD

Network models of brain disorders also suggest that spatially distributed structural alterations may reflect the connectivity profiles of specific regions that act as candidate epicenters.^26,37,41^ Regions whose normative functional or structural connectivity profiles showed significant spatial correlations with the regional morphometric difference maps were considered epicenters (**Fig. 4A**). We identified functional cortical epicenters, forming a distributed network involving medial temporal and orbitofrontal regions, with additional extensions into frontoparietal cortices, including the lateral orbitofrontal, entorhinal, inferior temporal, and banks of the superior temporal sulcus (**Fig. 4B**; Supplementary **Table 14**). Structural cortical epicenters were more circumscribed, with the strongest associations observed in sensorimotor and frontoparietal regions, including the precentral gyrus, inferior frontal gyrus (pars opercularis), caudal middle frontal gyrus, and inferior parietal cortex (**Fig. 4B**; Supplementary **Table 14**). Subcortical epicenters primarily involved the hippocampus, amygdala, and striatal regions (**Fig. 4B**; Supplementary **Table 15**).

**Fig. 4.**
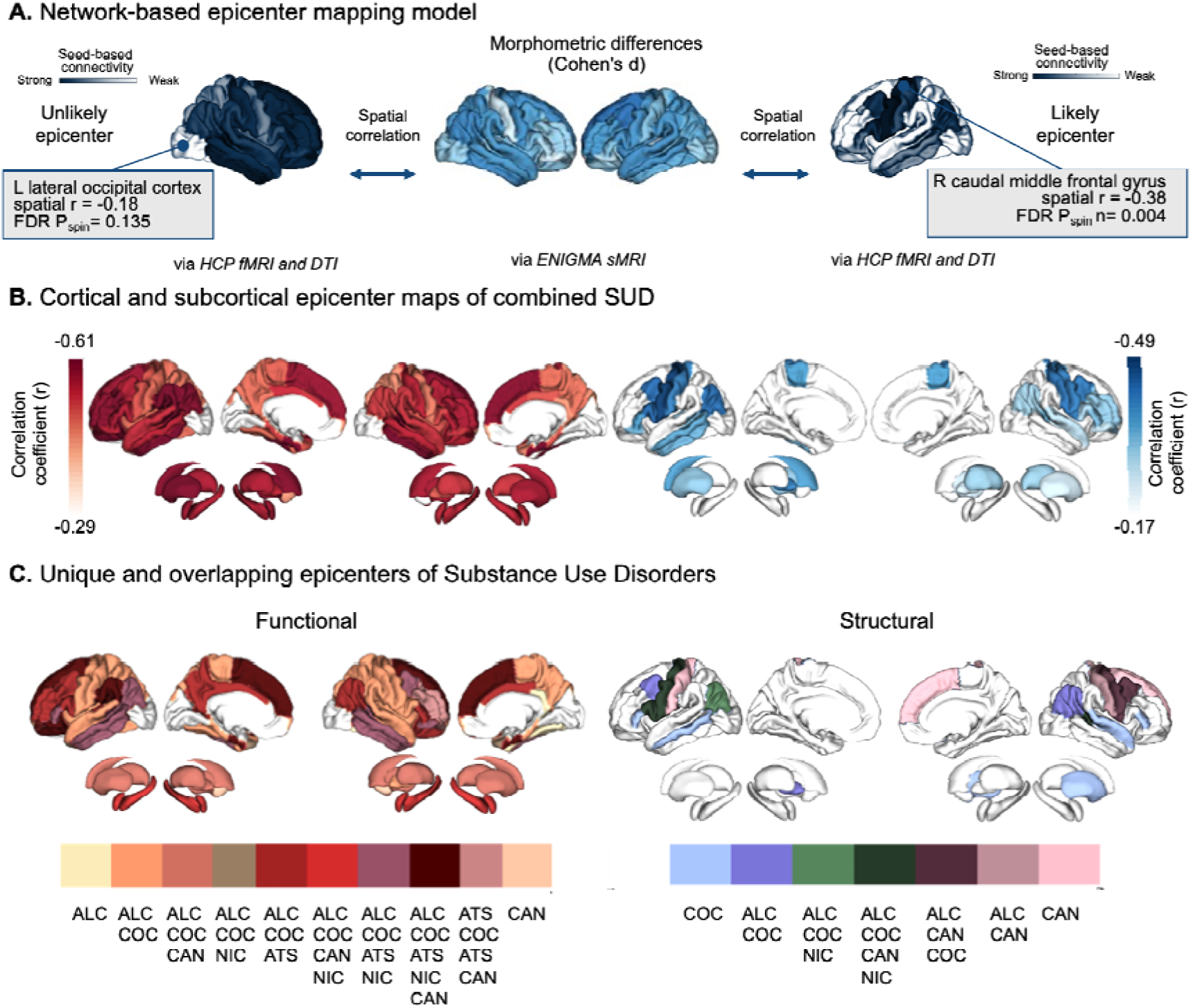
Functional and structural epicenters in SUD. (**A**) Model of network-based epicenter mapping. Spatial correlation between normative connectivity profiles and observed SUD-related cortical differences is used to identify likely versus unlikely epicenters. (**B**) Cortical and subcortical epicenter maps of the combined SUD pattern, derived from spatial correlations between normative functional (left) or structural (right) connectivity and cortical thickness differences in SUD. Darker colours indicate higher epicenter likelihood (Pearson’s *r*). Only regions showing significant spatial correlation with the maps of morphometric difference are displayed as putative epicenters (FDR-*P*_spin_ < 0.05 for cortical regions; FDR-P_shuff_ < 0.05 for subcortical regions; 10,000 permutations). (**C**) Overlap of functional and structural epicenters across substances. Shared epicenters are visualized as darker (more extensive overlap) and highlight regions which are shared epicenters, whereas unique epicenters are visualized lighter and highlight substance-specific epicenters. Only epicenters surviving multiple-comparison correction are displayed (FDR-Pspin < 0.05 for cortical regions; FDR-P_shuff_ < 0.05 for subcortical regions). Abbreviations: ALC = alcohol use disorder; ATS = amphetamine-type stimulant use disorder; CAN = cannabis use disorder; COC = cocaine use disorder; DTI = diffusion tensor imaging; ENIGMA = Enhancing NeuroImaging Genetics through Meta-Analysis; FDR = false discovery rate; fMRI = functional MRI; HCP = Human Connectome Project; NIC = nicotine use disorder; sMRI = structural MRI; SUD = substance use disorder. Cohen’s d = standardized mean difference; *P*spin = spin-permutation *P*-value; P_shuff_ = label-shuffling P-value; *r* = Pearson correlation coefficient.

### Similarity structure of substance-specific epicenter maps

Using substance-specific Cohen’s *d* maps, we identified epicenters for each single SUD (Supplementary **Fig. 3**) and quantified their spatial similarity to the combined SUD epicenter profile. Cortical epicenter maps showed high concordance with the combined SUD epicenters across substances, particularly for alcohol and cocaine (all *r* > 0.82; FDR-*P*_spin_ < 0.05; Supplementary **Table 16**). Cross-correlation analyses revealed greater variability across substances at the subcortical level (Supplementary **Table 17**). Given this high overall concordance, we examined the overlap and uniqueness of epicenters across substances (**Fig. 4C**). Across functional and structural cortical maps, epicenters shared by multiple substances largely overlapped with regions showing the highest epicenter likelihood in the combined SUD map (**Fig. 4B**), including frontal, parietal, and lateral temporal association areas. Substance-specific epicenters were comparatively sparse and modality-dependent, with distinct cortical loci for alcohol, cocaine, and cannabis and limited subcortical specificity (**Fig. 4C**; Supplementary **Tables 18–19**). Together, these results reveal a core set of frontal, parietal, and temporal epicenters anchoring cortical vulnerability across substances, alongside more circumscribed substance-specific loci reflecting variation in network-constrained morphometric differences. Further details are provided in the Supplementary Results.

### Robustness and generalizability analyses

Robustness and generalizability analyses included age and sex interaction, individual-level variability, age distribution, sample size, regional pattern stability, and control-sample composition. SUD-by-age interactions were cortical but not subcortical, and followed hub vulnerability and epicenter patterns similar to the main SUD morphometric maps (**Fig. 2C**; Supplementary **Fig. 4**; Supplementary **Tables 20–22**). SUD-by-sex interactions were absent cortically and limited subcortically to bilateral putamen and thalamus (**Fig. 2D**; Supplementary **Tables 23–24**). Individual-level analyses showed consistent but less sensitive hub vulnerability and epicenter frequency patterns (Supplementary **Fig. 5**; Supplementary **Table 25**). Age-harmonized resampling showed that larger alcohol and cocaine effects in the primary sample were not explained solely by older age distributions or SUD-by-age interaction effects (**Fig. 2A**; Supplementary **Fig. 6**; Supplementary **Tables 26–27**). Sample-size equalization, spatial stability analyses, and alternative control-sampling strategies supported stable substance-specific effect magnitudes and regional morphometric patterns (Supplementary **Figs 7-9**; Supplementary **Tables 28–30**). Additional details are provided in the Supplement.

### Transdiagnostic disorder comparison

Having established the robustness of the SUD epicenter architecture, we next examined the extent to which these patterns are shared across major psychiatric disorders, comparing SUD epicenters with those of six disorders using ENIGMA meta-analytic maps. Meta-analytic case-control maps from six major psychiatric disorders were used to generate epicenter maps for schizophrenia,^58,59^ bipolar disorder,^60,61^ major depressive disorder,^62,63^ obsessive-compulsive disorder,^64,65^ attention-deficit/hyperactivity disorder,^66,67^ and autism spectrum disorder,^68^ (Supplementary **Tables 31–34**).

Across cortical and subcortical domains, SUD epicenters showed strongest spatial correspondence with schizophrenia and bipolar disorder in both functional and structural modalities (*r* = 0.55–0.87, *P*_spin_ ≤ 0.047; **Fig. 5A**; Supplementary **Tables 35–36**). Correlations with obsessive-compulsive disorder, attention-deficit/hyperactivity disorder, and autism spectrum disorder were comparatively weaker. Major depressive disorder showed a distinct pattern, in which stronger subcortical functional epicenter expression in substance use disorders corresponded to weaker expression in major depressive disorder (*r* = −0.71, *P*_spin_ = 0.011; Supplementary **Table 36**). Substance-specific epicenter profiles largely recapitulated these transdiagnostic trends, while also revealing substance-dependent deviations, most prominently for opioids, which showed particularly strong correspondence with major depressive disorder. To further characterize the anatomical basis of this convergence, we examined regional epicenters. Functional epicenters were predominantly shared between SUD, schizophrenia, and bipolar disorder, involving frontotemporal and parietal cortices, as well as bilateral caudate nuclei and the left putamen (**Fig. 5B**; Supplementary **Tables 37–38**). A limited number of regions, including the left frontal pole and right temporal pole, also overlapped with major depressive disorder. In contrast, overlap of structural epicenters was sparse and largely restricted to frontoparietal cortical regions shared between SUD, schizophrenia, and bipolar disorder. No cortical or subcortical epicenter overlap was observed with obsessive-compulsive disorder, attention-deficit/hyperactivity disorder, or autism spectrum disorder (**Fig. 5B**; Supplementary **Tables 37–38**).

**Fig. 5.**
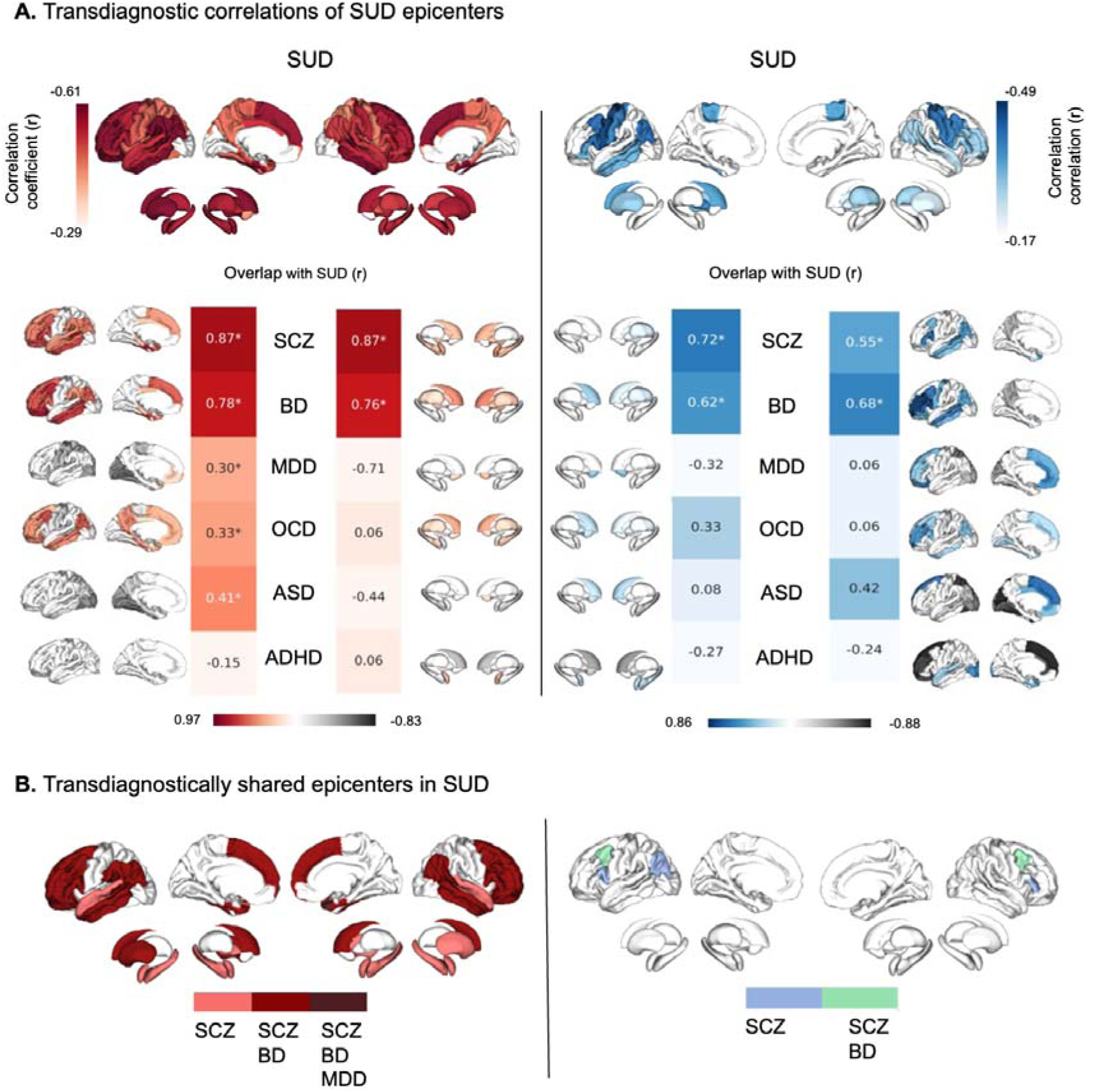
Correlations between SUD and psychiatric disorders. (**A**) Top: SUD epicenter map. Bottom: Spatial correlations of SUD cortical and subcortical epicenter maps and each psychiatric disorder (schizophrenia, bipolar disorder, major depressive disorder, obsessive-compulsive disorder, attention-deficit/hyperactivity disorder, and autism spectrum disorder), shown separately for functional and structural modalities. For visualization purposes, epicenter maps for the remaining psychiatric disorders are shown without multiple-testing correction (*P*_spin_ < 0.05), to illustrate overall pattern similarity rather than individual epicenters. (**B**) Brain map of shared and disorder-specific epicenters across SUD and the six psychiatric disorders, showing overlap in both functional and structural connectivity. Shared and unique epicenters represent regions surviving multiple-testing correction (FDR-*P*_spin_ < 0.05). Abbreviations: ADHD = attention-deficit/hyperactivity disorder; ASD = autism spectrum disorder; BD = bipolar disorder; MDD = major depressive disorder; OCD = obsessive-compulsive disorder; *r* = Pearson correlation coefficient; SCZ = schizophrenia; SUD = substance use disorder. Asterisks indicate significant correlations after spin-permutation testing and false discovery rate correction.

### Association of neurotransmitter density with cortical differences in SUD

Endogenous neurotransmitter systems are key mediators of exogenous substance effects^6,35^ and are closely linked to large-scale brain network architecture.^44,45^ Building on evidence for network-level involvement in SUD, we examined the relationship between neurotransmitter receptor distribution, SUD-related cortical differences, and intrinsic connectivity organization. We applied partial least squares (PLS) analysis to relate regional neurotransmitter receptor density profiles^44^ (Supplementary **Table 39**) to the patterns of substance-specific cortical thickness differences (**Fig. 2E**; Supplementary **Table 4**). The decomposition revealed four statistically significant latent variables (LVs; *P*_spin_ < 0.05, **Fig. 6A**), with the first two explaining over 90% of the cross-covariance (LV1 = 75.5%, LV2 = 15.9%; **Fig. 6B**). Both LV1 and LV2 aligned with the distributed cortical difference pattern (**Fig. 6C**). LV1 emphasized frontotemporal vulnerability, whereas LV2 additionally captured parietal involvement; both patterns correlated strongly with the combined SUD morphometric difference map (LV1 *r* = 0.97, *P*_spin_ < 0.001; LV2 *r* = 0.86, *P*_spin_ < 0.001). The primary axis of covariance (LV1; 75.5%) indicated that regions with the greatest morphometric differences exhibited higher normative density of κ-opioid receptor (KOR), metabotropic glutamate receptor 5 (mGluR5), cannabinoid receptor type 1 (CB1), histamine H3 receptor (H3), and μ-opioid receptor (MOR) markers (all loading z-scores > 1.96; **Fig. 6D)**. The second axis (LV2; 15.9%) indicated that these regions were, to a lesser extent, characterized by lower normative density of dopamine D1 receptor (D1), vesicular acetylcholine transporter (VAChT), dopamine transporter (DAT), and N-methyl-D-aspartate receptor (NMDA) markers (all loading *z*-scores < −1.96; Fig. 6D). Importantly, both LVs were related to normative connectome architecture. Lower LV1 and LV2 scores, indicating lower cortical thickness in SUD, were associated with higher functional (LV1 *r* = −0.37, *P*_spin_ = 0.017; LV2 *r* = −0.51, *P*_spin_ = 0.001) and structural centrality (LV1 *r* = −0.43, *P*_spin_ < 0.001; LV2 *r* = −0.48, *P*_spin_ < 0.001; **Fig. 6E).**

**Fig. 6.**
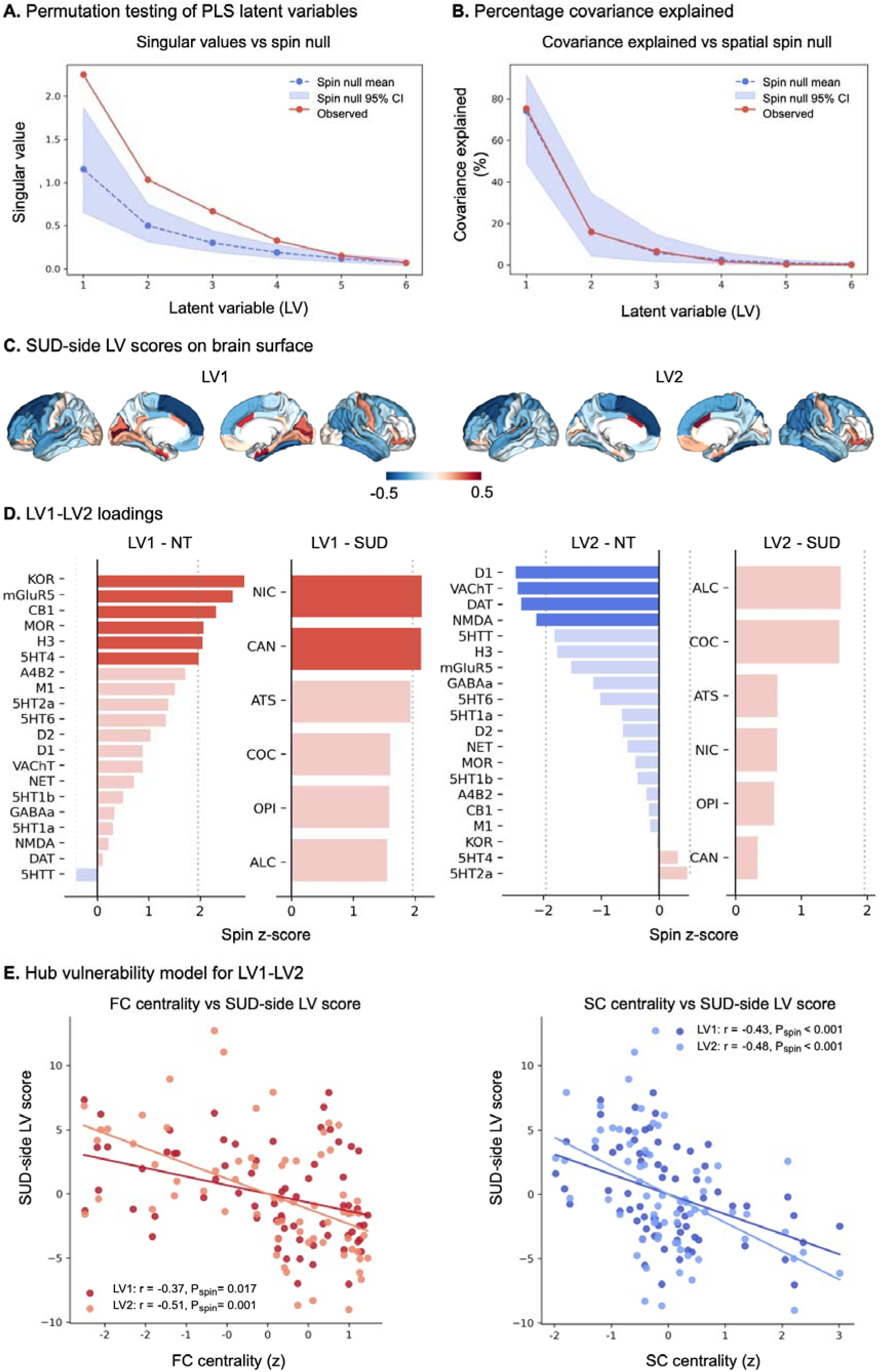
Association of neurotransmitter density with cortical differences in SUD. (**A**) Significance testing of partial least squares (PLS) latent variables (LVs), comparing observed singular values with a spatial spin-permutation null distribution (mean and 95% CI). (**B**) Covariance explained by each LV, shown relative to a spatial spin-permutation null distribution, with LV1 and LV2 jointly accounting for ∼90% of the total cross-covariance between standardized neurotransmitter receptor density and SUD-related cortical thickness differences. (**C**) Cortical maps of LV1 and LV2 SUD-side LV scores, computed as the projection of the SUD morphometric maps onto each latent variable, illustrating a shared frontotemporal vulnerability axis (LV1) and a secondary pattern with additional parietal involvement (LV2). By the sign convention used here, lower SUD-side LV scores (blue) correspond to regions with lower cortical thickness in SUD. (**D**) Variable loadings for each neurotransmitter receptor and each substance map on LV1 and LV2, expressed as *z*-scores relative to the spatial spin-permutation null distribution. On the neurotransmitter side, the largest positive LV1 loading *z*-scores were observed for KOR, mGluR5, CB1, MOR and H3, whereas nicotine and cannabis showed the largest positive SUD-side loading *z*-scores. For LV2, D1, VAChT, DAT and NMDA showed the largest negative neurotransmitter-side loading *z*-scores, whereas alcohol and cocaine showed the largest positive SUD-side loading *z*-scores, although the latter did not reach the significance threshold. (**E**) Associations between SUD-side LV scores and cortical degree centrality. Functional centrality was negatively associated with both LV1 (*r* = −0.37, *P*_spin_ = 0.017) and LV2 (*r* = −0.51, *P*_spin_ = 0.001) SUD-side scores; corresponding negative associations were observed for structural centrality (LV1 *r* = −0.43, *P*_spin_ < 0.001; LV2 *r* = −0.48, *P*_spin_ < 0.001). Thus, lower SUD-side LV scores, indicating lower cortical thickness in SUD, were associated with higher functional (FC) and structural (SC) degree centrality, consistent with preferential involvement of highly connected cortical hubs. Abbreviations: A4B2 = α4β2 nicotinic acetylcholine receptor; ALC = alcohol use disorder; ATS = amphetamine-type stimulant use disorder; CAN = cannabis use disorder; CB1 = cannabinoid receptor type 1; CI = confidence interval; COC = cocaine use disorder; D1 = dopamine D1 receptor; D2 = dopamine D2 receptor; DAT = dopamine transporter; FC = functional connectivity; GABAa = γ-aminobutyric acid type A receptor; H3 = histamine H3 receptor; KOR = κ-opioid receptor; LV = latent variable; mGluR5 = metabotropic glutamate receptor 5; MOR = μ-opioid receptor; NET = norepinephrine transporter; NIC = nicotine use disorder; NMDA = N-methyl-D-aspartate receptor; NT = neurotransmitter; OPI = opioid use disorder; *P*spin = spin-permutation *P*-value; *r* = Pearson correlation coefficient; SC = structural connectivity; SUD = substance use disorder; VAChT = vesicular acetylcholine transporter; 5HT = serotonin (5-hydroxytryptamine), with receptor labels indicating serotonin receptor subtypes.

## Discussion

Leveraging the largest structural neuroimaging cohort in addiction research to date, this study establishes the network and neurochemical principles governing brain-wide structural vulnerability in SUD. Across the six substance use disorders, we identified a reproducible, system-wide pattern of cortical and subcortical differences involving frontal, parietal and temporal association cortex and limbic subcortical structures. Our findings align with previous results^8,70–74^ and extend them by revealing broader regional involvement in SUD pathology. Significant SUD-by-age interactions indicated steeper age-related cortical thinning in SUD compared to controls, consistent with cumulative exposure-related effects and increased vulnerability to age-related brain changes. Sex interaction analyses revealed only subtle, subcortically-restricted differences, suggesting the main morphometric signature is broadly consistent across biological sexes. Across substances, we observed high spatial similarity, consistent with shared effects in distinct SUD.^8,70,71^ Effect sizes were largest for alcohol and cocaine, and differed across substances. Overall, structural alterations in SUD were system-wide, graded across substances, and comparable in magnitude to those reported for schizophrenia and bipolar disorder.^58–61^

In line with the hub vulnerability hypothesis,^29,75^ cortical morphometric differences in SUD were associated with functional and structural degree centrality, with greater alterations in highly connected hub regions. Cortical hubs are characterized by protracted developmental plasticity,^76,77^ dense neurotransmitter receptor expression,^44,45^ and high metabolic demand.^32,78^ They may therefore be particularly vulnerable to excitotoxicity, oxidative stress and inflammatory processes,^79^ especially in chronic substance exposure.^80,81^ Chronic stimulation of receptor systems concentrated in the association cortices may, through converging molecular pathways, contribute to the observed lower cortical thickness and age-dependent effects in hub regions.

We observed shared and substance-specific hub vulnerability patterns, with alcohol and cocaine showing stronger associations with functional hubs, whereas cannabis and nicotine showed relatively stronger associations with structural hubs. This divergence may reflect differences between functional connectivity, capturing transient co-activation, and structural connectivity, reflecting more stable anatomical pathways.^30,82^ Such differences may begin to explain why certain substances preferentially affect functional coupling, whereas others alter structural integrity.^13,79^ Future studies are necessary to clarify whether these differences are driven by substance-specific pharmacodynamic properties or heterogeneity in substance-use patterns.

Epicenter mapping identified regions whose normative connectivity profiles matched the spatial distribution of SUD-related morphometric alterations, with functional epicenters distributed across associative networks and structural epicenters more confined to unimodal and sensorimotor territories. This suggests that SUD-related alterations may arise from multiple spatial “seeds” rather than a single focal point, consistent with parallel involvement of multiple neural circuits during chronic substance exposure.^21,35,36^ Multiple epicenters may reflect combined distributed substance-related neurotoxic effects and neuroadaptation,^21,35,83^ as well as pre-existing network vulnerabilities^30,57^ rather than a single cascade. Compared to other neuropsychiatric disorders,^25,28^ epicenters in SUD appeared more diffuse, potentially reflecting cumulative effects of substance exposure, including neuroadaptation as well as broader brain health processes such as neuroinflammation and cardiovascular burden.^22,84–87^ In a transdiagnostic context, SUD epicenters overlapped substantially with those of schizophrenia and bipolar disorder,^26^ particularly within salience and frontoparietal control networks.^88,89^ Structural epicenter overlap was more restricted but localized to inferior parietal cortex and inferior frontal gyrus (pars opercularis), key nodes of frontoparietal control and salience/inhibitory networks.^90^ These networks support interoception, attention, salience attribution and reward,^89,91^ functions affected across SUD, schizophrenia, and bipolar disorder and potentially representing shared mechanisms of vulnerability.^92^ Although schizophrenia and bipolar cohorts show high rates of nicotine dependence and comorbid SUD,^93–97^ this concern is mitigated by exclusion of major psychiatric disorders in our SUD cohorts and of current non-nicotine SUD in most schizophrenia and bipolar neuroimaging studies.^58–61^

Our multivariate neurotransmitter mapping indicated that SUD-related morphometric differences align along two principal axes of neurotransmitter architecture. The dominant latent variable (LV1) loaded on κ-opioid, µ-opioid, mGluR5, CB1, and H3 receptors and captured the cortical signature of the combined SUD phenotype, particularly in frontotemporal association cortices. This is consistent with prior work showing that transmodal association cortex and hub-like regions are enriched in multiple receptor classes and anchor cross-disorder vulnerability gradients.^45,98–101^ A second weaker latent variable (LV2) showed greater parietal involvement and was associated with lower normative densities of D1, DAT, VAChT, and NMDA. This axis may reflect secondary vulnerability of regions with relatively sparse neuromodulatory input, potentially conferring reduced resilience to widespread neurochemical perturbations.^102–104^ Dopamine and glutamate signaling are implicated in neurotrophic and spine-stabilizing processes,^105–109^ and disruptions during chronic substance use have been linked to synaptic and dendritic alterations in association circuits.^4,110–112^ Notably, the spatial expression of both axes was associated with cortical hub topology, such that regions showing greater LV-related morphometric difference also showed higher functional and structural degree centrality. It remains an open question whether regional receptor density predisposes areas to structural change, whether morphometric alterations secondarily affect receptor expression, or whether both reflect shared principles of network organization and chemoarchitecture.^56,98^

Our study has limitations. Although this represents the largest SUD neuroimaging cohort to date, sample sizes for opioids and amphetamine-type stimulants were modest and findings for these groups should be interpreted cautiously. Clinical heterogeneity and polysubstance use may have obscured morphometric brain differences unique to each substance. The cross-sectional design precludes causal inference and cannot disentangle temporary neuroplastic adaptations from longer-lasting neurotoxic effects, nor account for variation in substance exposure trajectories, consumption levels, or abstinence patterns. Harmonized clinical measures of severity and exposure were also not uniformly available across the 51 contributing sites, precluding analyses linking morphometric differences to clinical variation. Longitudinal MRI studies demonstrate partial structural recovery with decreased consumption or abstinence,^17,19^ while animal models show that neurodegeneration and regeneration can co-occur, as a function of intake and abstinence duration.^113,114^

In sum, addiction-related brain morphometric differences are shaped by connectome organization and neurotransmitter receptor architecture. Across six SUD, we identified a largely shared pattern of cortical and subcortical differences, age-dependent cortical vulnerability, preferential involvement of cortical hubs, distributed epicenters, transdiagnostic overlap with schizophrenia and bipolar disorder, and receptor-defined neurochemical axes. These findings suggest that normative connectivity and chemoarchitecture constitute organizing constraints shaping SUD-related brain pathology. Together, this systems-level framework supports the identification of receptor- and circuit-based targets for intervention in SUD.

## Supporting information

Supplementary Methods and Results

## Data availability

All data reported in the manuscript are included in the article and its supplementary information files. Summary statistics of the case-control meta-analyses from the ENIGMA Working Groups are available from the ENIGMA Toolbox (https://enigma-toolbox.readthedocs.io/en/latest/).^46^ Preprocessed normative cortico-cortical, subcortico-cortical, and subcortico-subcortical functional and structural connectivity data are available from the ENIGMA Toolbox (https://enigma-toolbox.readthedocs.io/en/latest/). Molecular, connectivity, and receptor datasets are available from https://github.com/netneurolab/hansen_crossdisorder_vulnerability.git and https://github.com/netneurolab/hansen_receptors.git. Additional information can be made available upon reasonable request to the authors.

## Code availability

All software and analysis code used in this study are openly available. Image preprocessing pipelines are available through the ENIGMA consortium (https://github.com/ENIGMA-git). The network modelling code is publicly available from the ENIGMA toolbox https://enigma-toolbox.readthedocs.io/en/latest.^46^ All further statistical analyses were performed in R version 4.3.0. The respective packages are mentioned in the corresponding “Methods” sections.

## Acknowledgements

The authors used Claude (Opus 4.8 and Opus 5; Anthropic) and ChatGPT (GPT-4o and GPT-5-series models; OpenAI) to check the manuscript for spelling and grammatical errors, formatting consistency, and clarity of language. No text, data, figures, or interpretations were generated by these tools. All output was reviewed by the authors, who take full responsibility for the content of this article.

## Supplementary material

Supplementary material is available online with the article.

