## Supplementary Methods and Results for "Network and receptor architectures shape brain morphometry in addiction"

Georgiadis & Milano et al.

**Supplementary Methods and Results**

### Site selection and demographics

Inclusion and exclusion criteria for each site, and participant demographics (age, sex) for healthy controls and six primary substance use disorder (SUD) groups (alcohol, amphetamines, cannabis, cocaine, nicotine, opioids) are listed in Supplementary **Table 1**. Across all 51 sites, substance use disorder diagnosis was confirmed using Diagnostic and Statistical Manual of Mental Disorders, Fourth Edition (DSM-IV), or International Classification of Diseases, 10th Revision (ICD-10), criteria. The geographic distribution of participating Enhancing NeuroImaging Genetics through Meta-Analysis (ENIGMA) Addiction Working Group cohorts is shown in Supplementary **Fig. 1**.

The pooled sample included 1,951 healthy controls (mean age ± standard deviation [SD] = 33.8 ± 12.0 years; 37.5% female) and 2,782 individuals with SUD (mean age ± SD = 34.6 ± 11.2 years; 33.3% female). Of the SUD group, 2,312 participants (mean age ± SD = 34.7 ± 11.3 years; 33.9% female) met criteria for a single SUD diagnosis without comorbidity and were included in substance-specific analyses (alcohol: 902; amphetamines: 178; cannabis: 286; cocaine: 278; nicotine: 600; opioids: 68; Supplementary **Table 2**).

#### Fig. 1. Worldwide distribution of ENIGMA Addiction Working Group Cohorts

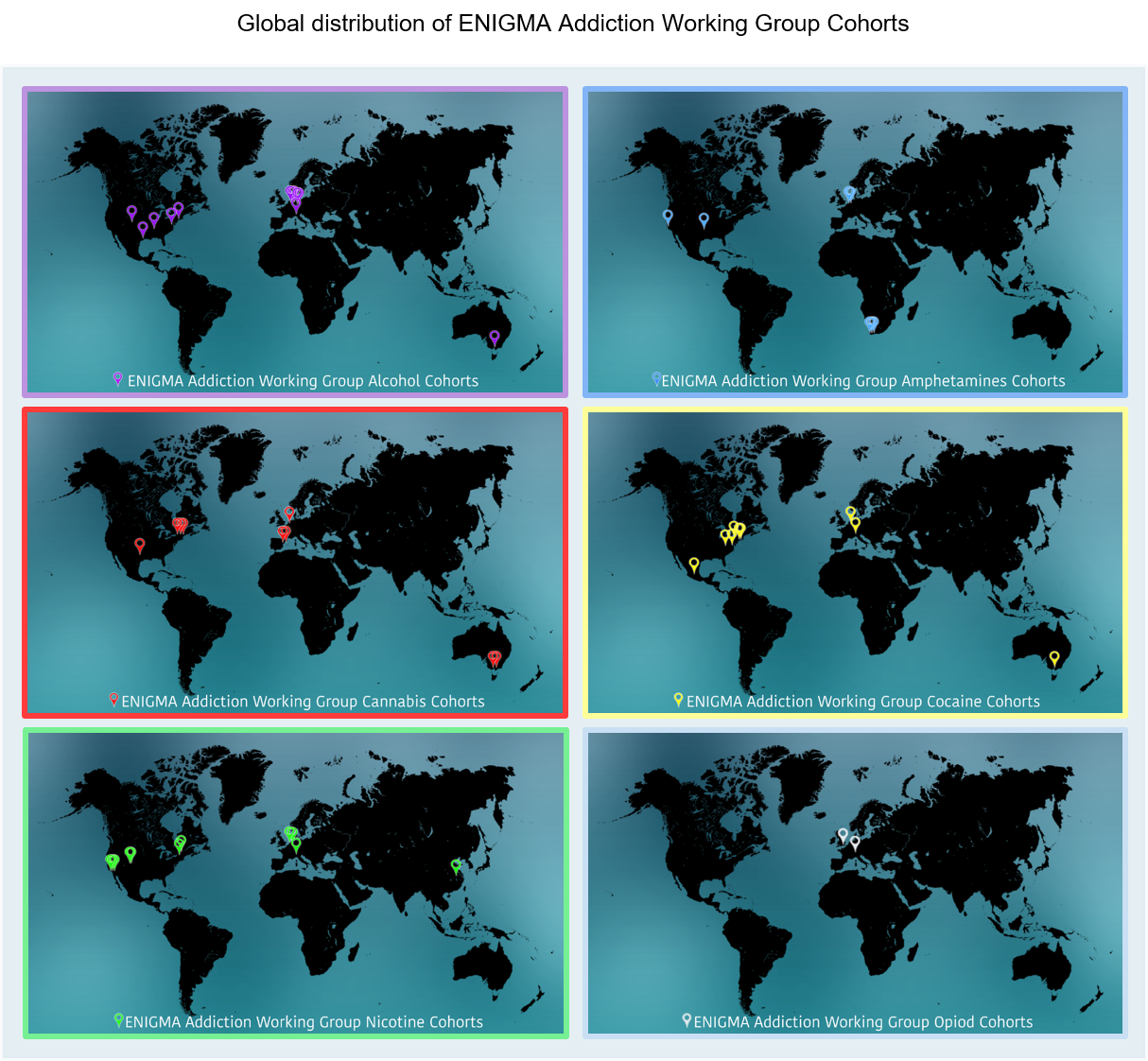

**Supplementary Fig. 1.** World maps illustrate the geographic locations of participating sites contributing structural MRI data to the ENIGMA Addiction Working Group across six primary substance use disorder categories: alcohol, amphetamines, cannabis, cocaine, nicotine and opioids. Each panel corresponds to one substance-specific cohort.

Abbreviations: ENIGMA = Enhancing NeuroImaging Genetics through Meta-Analysis.

**Alt text:** World maps showing the geographic distribution of ENIGMA Addiction Working Group cohorts contributing structural MRI data across alcohol, amphetamine, cannabis, cocaine, nicotine, and opioid use disorder groups.

### MRI acquisition and preprocessing across cohorts

T1-weighted structural MRI scans were acquired using magnetization-prepared rapid gradient-echo (MPRAGE) sequences across cohorts, with additional use of gradient echo, multi-echo MPRAGE (MEMPRAGE), and other structural protocols at specific sites. Acquisition parameters varied across sites but typically involved repetition times (TR) ranging from ~1900 to 2530 ms, with shorter TRs used in selected alternative protocols, and echo times (TE) between ~1.5 and 5 ms. Voxel sizes were generally isotropic at 1×1×1 mm³, although some cohorts employed non-isotropic or submillimetric resolutions (e.g., 0.9×0.9×0.9 mm³, 1.2×1.2×1.2 mm³, or 0.47×0.47×1 mm³), with matrix sizes commonly around 256×256×176 but reaching higher in-plane resolutions at selected sites.

Flip angles for MPRAGE sequences ranged from 6° to 15° across cohorts, with higher values observed in specific T1-weighted protocols, such as the Gradient Echo Volumetric sequence in the nicotine cohort (35°). Functional echo-planar imaging (EPI) acquisitions in the nicotine cohort used higher flip angles (82°), consistent with standard functional MRI (fMRI) protocols. In addition to standard MPRAGE, sites employed alternative T1-weighted sequences, including MEMPRAGE for alcohol; spoiled gradient-recalled echo (SPGR) for amphetamines and cannabis; Fast Spoiled Inversion-Recovery for cannabis; brain volume imaging (BRAVO) for alcohol; Turbo Field Echo for cocaine and cannabis; and fast spoiled gradient-recalled echo (FSPGR) for nicotine, each with distinct acquisition characteristics. Some cohorts, such as cannabis, included high-resolution scans with submillimetric voxel dimensions (e.g., 0.47×0.47×1 mm³).

Following image preprocessing, all structural data were parcellated using the Desikan–Killiany atlas, yielding 68 cortical and 14 subcortical regions.^1^ Despite variability in scanner type and sequence implementation, acquisition protocols across cohorts ensured consistently high spatial resolution suitable for robust cortical and subcortical morphometric analyses (Supplementary **Table 3**).

### Normative connectivity construction

Normative functional and structural connectivity matrices were obtained from the ENIGMA Toolbox,^2^  based on resting-state fMRI and diffusion MRI data from 207 unrelated healthy young adults (83 males; mean age ± SD = 28.7 ± 3.7 years) from the Human Connectome Project (HCP). These data were preprocessed using the HCP minimal processing pipeline.^3,4^ The derivation of the matrices is described in detail in the ENIGMA Toolbox documentation.^2^ Following preprocessing and parcellation to the Desikan–Killiany atlas (68 cortical and 14 subcortical regions),^1^ individual functional and structural connectomes were computed. Functional connectomes were derived by computing Pearson correlations between the time series of each unique pairing of the 68 cortical regions (cortico-cortical connectivity) and each unique pairing of 14 subcortical to 68 cortical regions (subcortico-cortical connectivity). Negative values were set to zero, and after Fisher z-transformation, the matrices were averaged across subjects. Structural connectomes were generated using MRtrix3^5^ with anatomically constrained tractography,^6^ multi-tissue constrained spherical deconvolution,^7^ and spherical-deconvolution informed filtering of tractograms 2 (SIFT2)^8^ following constrained spherical deconvolution (CSD)-based fibre orientation estimation.^9^

Whole-brain tractography produced 40 million streamlines (maximum length = 250 mm; fractional anisotropy cutoff = 0.06), which were mapped to the 68x68 cortico-cortical and 14x68 subcortico-cortical matrices. Group-level structural connectivity matrices were obtained by applying distance-dependent thresholding^10^ and logarithmic transformation.

Based on these matrices, functional and structural cortico-cortical and subcortico-cortical weighted degree centrality were computed for each region. Formally, for node i in a weighted network, degree centrality was defined as:

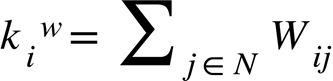

where *N* is the set of all nodes in the network and W*ij* represents the weight of the connection between nodes *i* and *j.* For each brain region, weighted cortico-cortical and subcortico-cortical degree centrality was calculated as the sum of all connection weights to other regions, excluding self-connections^11^.

Group-average matrices exhibited canonical hub topology, with highest centrality in medial prefrontal regions.

### Batch-effect correction

ComBat harmonization was applied to cortical thickness and subcortical volumes to mitigate site-related variance.^12,13^ No partial R² analysis was performed, as effect sizes were summarized using Cohen’s *d*.

### Structural differences across SUD

We compared cortical thickness and subcortical volumes between individuals with substance use disorders and healthy controls using region-wise linear regression models that included diagnostic group, age, sex, and intracranial volume (ICV) as covariates for both cortical and subcortical measures. In the cortical analyses, we assessed thickness across the 68 regions of the Desikan–Killiany atlas, extracting *T*-statistics, false discovery rate (FDR)-corrected *P*-values, and Cohen’s *d* effect sizes (Supplementary **Table 4**). Similarly, subcortical volumes were analyzed for 14 subcortical structures: bilateral nuclei, namely, the nucleus accumbens, amygdala, caudate, pallidum, putamen, thalamus, and hippocampus, using the same statistical approach (Supplementary **Table 5**). We report results both for the entire cohort of individuals with substance use disorders and for each specific substance: alcohol, amphetamines, cannabis, cocaine, nicotine, and opioids.

In the overall substance use disorder group, 65 out of 68 cortical regions showed significantly reduced thickness compared to controls (FDR- corrected *P*  < 0.05). The most pronounced effects were observed in prefrontal and parietal regions, including the left superior frontal gyrus (*T* = –11.36; Cohen’s *d*  = –0.33), the left caudal middle frontal gyrus (*T* = –10.41; Cohen’s *d*  = –0.31), bilateral superior parietal cortices (left: *T* = –9.50, Cohen’s *d* = –0.28; right: *T* = –9.66, *d* = –0.28) and the left precuneus (*T* = –9.05; Cohen’s *d*  = –0.27). Subcortical analyses revealed significant volume reductions in 11 out of 14 regions. In particular, in the hippocampus (left: *T* = –8.39, *d* = –0.25; right: *T* = –8.31, *d* = –0.24), amygdala (left: *T* = –8.05, *d* = –0.24; right: *T* = –7.10, *d* = –0.21), and thalamus bilaterally (left: *T* = –5.01, *d* = –0.15; right: *T* = –4.97, *d* = –0.15). Smaller effects were observed in the caudate (left: *d* = –0.10; right: *d* = –0.08), and more heterogeneous/lateralized effects in the accumbens (left: *d* = –0.04, non-significant, right: *d* = –0.11), pallidum (left: *d* = –0.05, non-significant, right: *d* = –0.12) and putamen (left: *d* = –0.06; right: *d* = –0.03, non-significant).

#### Sensitivity to non-linear age effects

Because cortical thickness and subcortical volumes can show modest non-linear (curvilinear) associations with age across adulthood, we performed a sensitivity analysis to test whether the case–control effect size pattern depended on assuming a strictly linear age effect. Specifically, we re-fit the primary models after adding a quadratic age term (age²) while keeping all other covariates and analytical choices identical to the main analysis.

Including age² had a negligible impact on the estimated case–control differences. The Cohen’s *d* effect-size vectors from the linear and quadratic models were almost perfectly correlated (cortical: *r* = 0.998, *P* < 0.0001; subcortical: *r* = 0.997, *P* < 0.0001), and absolute differences in effect-size magnitude were minimal (cortical thickness [CT]: root mean square deviation [RMSD] = 0.006; subcortical volume [SV]: RMSD = 0.007); see *Age and sex modulation of cortical thickness and subcortical volume in SUD* below).

#### Leave-one-out analysis

Having established substantial overlap across substance-specific morphometric patterns, we next assessed whether the combined-SUD phenotype was driven by any single substance. To this end, we conducted a leave-one-out analysis by sequentially excluding each of the six SUD subtypes and correlating the resulting Cohen’s *d* profiles with the full combined map (Supplementary **Tables 6–7**).

Cortical leave-one-out correlations were uniformly high, ranging from *r* = 0.944 to *r* = 0.999 (Supplementary **Table 8**), indicating that the spatial topography of cortical thickness differences remained highly stable across all iterations. Subcortical leave-one-out correlations showed a similarly robust pattern, ranging from *r* = 0.947 to *r* = 0.999 (Supplementary **Table 9**). Together, these analyses confirm that the overall spatial pattern of morphometric alterations observed in the combined-SUD phenotype is not driven by any single substance, but instead reflects a robust and shared neuroanatomical signature across SUD subtypes.

### SUD-specific structural differences

#### PCA of shared brain morphometric patterns across SUD

To further highlight the common variance structure of each SUD, we performed principal component analysis (PCA) on the six cortical and six subcortical morphometric difference maps (Supplementary **Fig. 2A** and **B**). The first principal component (PC1) explained 72.4% of cortical and 76.2% of subcortical variance across SUD-related morphometric differences. Both cortical and subcortical PC1 maps were highly correlated with the corresponding combined-SUD morphometric maps shown in **Fig. 2B** in the main manuscript (cortical: *r* = 0.98, *P*_spin_ < 0.001; subcortical: *r* = 0.96, *P*_spin_ < 0.001).

#### Fig. 2. Cross-correlation structure and shared morphological pattern across SUD

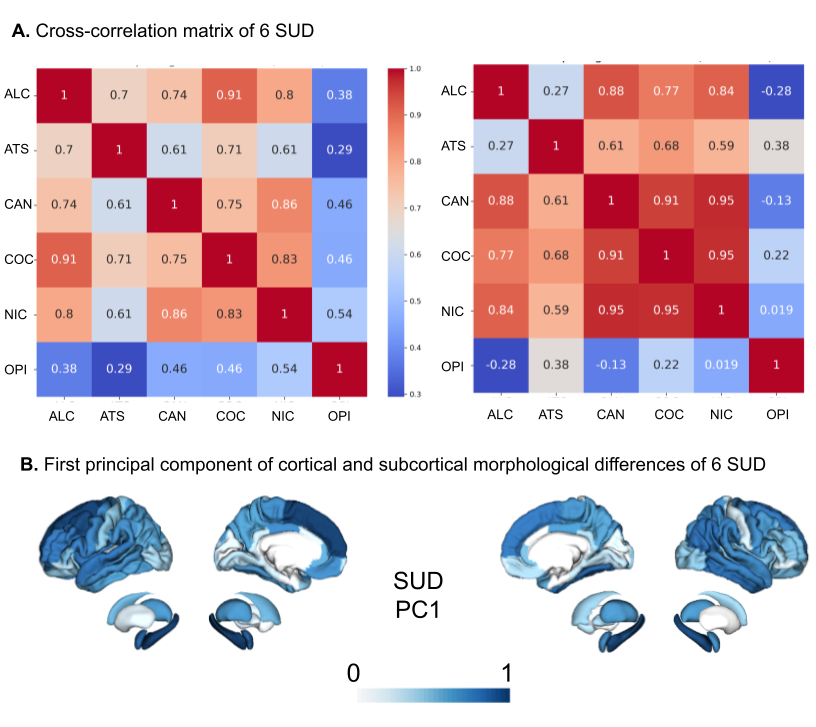

**Supplementary Fig. 2.** (**A**) Pairwise cross-correlation matrices showing similarity of cortical (left) and subcortical (right) morphometric difference maps across six substance use disorders. (**B**) Spatial projection of the first principal component (PC1) derived from cortical and subcortical morphometric differences, capturing the dominant shared pattern across substance use disorders.

Abbreviations: ALC = alcohol use disorder; ATS = amphetamine-type stimulant use disorder; CAN = cannabis use disorder; COC = cocaine use disorder; NIC = nicotine use disorder; OPI = opioid use disorder; PC1 = first principal component; PCA = principal component analysis; SUD = substance use disorder.

**Alt text:** Correlation matrices and brain maps showing shared cortical and subcortical morphometric patterns across substance use disorders, including the dominant principal component explaining common variance across substances.

### Substance-specific hub vulnerability

Hub vulnerability was assessed by correlating regional centrality in normative functional (resting-state) and structural (diffusion-based) networks with the spatial distribution of morphometric alterations in substance use disorders. For cortical analyses, centrality values were obtained for each of the 68 Desikan–Killiany regions, and significance was evaluated using 10,000 spin permutations, yielding spin-permutation P-values (*P*_spin_). For subcortical analyses, hub vulnerability was quantified using degree centrality in seven bilateral nuclei (nucleus accumbens, amygdala, caudate, hippocampus, pallidum, putamen, and thalamus), and significance was evaluated using label shuffling, yielding label-shuffling P-values (*P*_shuff_). Multiple-comparison correction was applied using the false discovery rate (FDR), and FDR-corrected *P*_spin_ or *P*_shuff_ values are reported where appropriate. Correlations with functional and structural centrality are denoted as r_func_ and r_struc_, respectively.

Across the combined SUD group, cortical regions with higher centrality showed significantly greater thinning both in functional (*r* = –0.51, *P*_spin_ = 0.002) and structural networks (*r* = –0.49, *P*_spin_ < 0.0001). At the substance-specific level, alcohol use disorder exhibited the strongest cortical hub vulnerability (r_func_ = –0.59, FDR-*P*_spin_ = 0.0006; r_struc_ = –0.52, FDR-*P*_spin_ = 0.0006). Amphetamine use disorder (r_func_ = –0.37, FDR-*P*_spin_ = 0.032; r_struc_ = –0.38, FDR-*P*_spin_ = 0.004) and cocaine use disorder (*r*_func_ = –0.51, FDR-*P*_spin_ = 0.012; r_struc_ = –0.47, FDR-*P*_spin_ = 0.003) also showed significant associations. In contrast, cannabis and nicotine use showed significant effects only in the structural network (cannabis: *r* = –0.30, FDR-*P*_spin_ = 0.012; nicotine: *r* = –0.35, FDR-*P*_spin_ = 0.006), while opioid use disorder did not show significant cortical hub vulnerability (r_func_ = 0.11, FDR-*P*_spin_ = 0.28; r_struc_ = –0.15, FDR-*P*_spin_ = 0.13) (Supplementary **Table 10**). At the subcortical level, the combined SUD group did not show significant hub vulnerability (r_func_ = –0.28, FDR-*P*_shuff_ = 0.16; r_struc_ = 0.21, FDR-*P*_shuff_ = 0.24). No substance subgroup exhibited significant subcortical hub vulnerability after FDR correction (Supplementary **Table 11**). To ensure robustness of our cortical hub vulnerability finding, we repeated the analysis across four centrality indices: degree, eigenvector, betweenness, and closeness. The centrality measures themselves showed strong inter-correlations within each modality (Supplementary **Table 12**) especially in structural networks (*r* = 0.787–0.983) with degree, eigenvector and closeness also highly correlated in functional networks (*r* = 0.960–0.996), while betweenness showed only moderate correlation**s** with the others (*r* = 0.345–0.476). Accordingly, we could confirm the consistency of hub vulnerability across these four centrality metrics in both functional and structural connectomes. The strongest and most consistent effects were observed in alcohol and cocaine use disorder. Amphetamine use disorder showed weaker but still significant associations across several metrics. By contrast, cannabis and nicotine use disorders displayed mostly non-significant or modest effects, with only a subset of structural metrics reaching significance. No significant hub vulnerability effects were detected for opioid use disorder (Supplementary **Table 13**).

### Combined SUD and substance-specific epicenter mapping

Epicenters were identified by correlating normative connectivity profiles with regional morphometric alterations (Supplementary **Fig. 3**). For cortical regions (Supplementary **Table 14**), we computed, for each of the 68 Desikan–Killiany parcels, the Pearson correlation between its normative functional (resting-state) or structural (diffusion-based) connectivity profile (derived from Human Connectome Project data) and the spatial map of cortical thickness alterations in substance use disorders. Subcortical epicenters (Supplementary **Table 15**) were derived analogously for seven bilateral nuclei (nucleus accumbens, amygdala, caudate, hippocampus, pallidum, putamen, and thalamus). Statistical significance was assessed using 10,000 *spin* permutations with FDR correction.

In the combined SUD cohort, functional cortical epicenters localized predominantly to limbic–temporal regions, with the strongest associations observed in the bilateral entorhinal cortex (left *r* = −0.61, FDR-*P*_spin_ < 0.0001; right *r* = −0.61, FDR-*P*_spin_ < 0.0001), bilateral inferior temporal gyrus (left *r* = −0.57, FDR-*P*_spin_ < 0.0001; right *r* = −0.51, FDR-*P*_spin_ < 0.0001), and banks of the superior temporal sulcus bilaterally (left *r* = −0.49, FDR-*P*_spin_ < 0.0001; right *r* = −0.49, FDR-*P*_spin_ = 0.001). Additional significant functional epicenters were detected in the left lateral orbitofrontal cortex (*r* = −0.52, FDR-*P*_spin_ = 0.001) and the right middle temporal gyrus (*r* = −0.41, FDR-*P*_spin_ < 0.0001).

Structural cortical epicenters were generally weaker and shifted toward fronto-parietal and sensorimotor regions, with peak associations in the left precentral gyrus (*r* = −0.49, FDR-*P*_spin_ < 0.0001), left pars opercularis (*r* = −0.42, FDR-*P*_spin_ = 0.002), left caudal middle frontal gyrus (r = −0.40, FDR-*P*_spin_ = 0.003), and left inferior parietal cortex (*r* = −0.40, FDR-*P*_spin_ = 0.002). Substance-specific analyses recapitulated these patterns. Alcohol showed strong functional cortical epicenters in the entorhinal cortex (left *r* = −0.64, FDR-*P*_spin_ < 0.0001; right *r* = −0.63, FDR-*P*_spin_ = 0.001) and inferior temporal gyrus (left *r* = −0.66, FDR-*P*_spin_ < 0.0001; right *r* = −0.58, FDR-*P*_spin_ < 0.0001), alongside a prominent structural epicenter in the left precentral gyrus (*r* = −0.49, FDR-*P*_spin_ = 0.001).

Amphetamines exhibited functional epicenters in limbic–temporal regions, including the left entorhinal cortex (*r* = −0.49, FDR-*P*_spin_ = 0.001) and left inferior temporal gyrus (*r* = −0.48, FDR-*P*_spin_ = 0.001), whereas structural epicenters were weaker and less spatially coherent, showing limited overlap with the functional pattern.

Cannabis showed functional epicenters in the entorhinal cortex bilaterally (left *r* = −0.38, FDR-*P*_spin_ = 0.004; right *r* = −0.41, FDR-*P*_spin_ = 0.003), with comparatively modest structural associations, most notably in the right caudal middle frontal gyrus (*r* = −0.38, FDR-*P*_spin_ = 0.006).

Cocaine demonstrated the most spatially extensive functional epicenter pattern, including banks of the superior temporal sulcus bilaterally (left *r* = −0.49, FDR-*P*_spin_ = 0.002; right *r* = −0.49, FDR-*P*_spin_ = 0.001), caudal middle frontal gyrus bilaterally (left r = −0.44, FDR-*P*_spin_ = 0.001; right r = −0.50, FDR-*P*_spin_ < 0.0001), inferior parietal cortex (left *r* = −0.50, FDR-*P*_spin_ < 0.0001), rostral middle frontal gyrus (left *r* = −0.46, FDR-*P*_spin_ < 0.0001), entorhinal cortex (left *r* = −0.62, FDR-*P*_spin_ < 0.0001; right *r* = −0.66, FDR-*P*_spin_ < 0.0001), and inferior temporal gyrus (left *r* = −0.61, FDR-*P*_spin_ = 0.001). Structural epicenters in cocaine were also evident but more circumscribed, with significant associations in the left inferior parietal cortex (r = −0.41, FDR-*P*_spin_ < 0.0001), left pars opercularis (*r* = −0.42, FDR-*P*_spin_ = 0.001), and left caudal middle frontal gyrus (*r* = −0.41, FDR-*P*_spin_ = 0.002).

Nicotine showed functional epicenters concentrated in the entorhinal cortex bilaterally (left r = −0.48, FDR-*P*_spin_ = 0.002; right r = −0.50, FDR-*P*_spin_ < 0.0001), with weaker and less consistent structural associations. Opioids exhibited comparatively weaker cortical epicenter effects; the most consistent finding was a functional epicenter in the right pars orbitalis (r = −0.31, FDR-*P*_spin_ = 0.017), which was also observed structurally (*r* = −0.24, FDR-*P*_spin_ = 0.025), alongside additional modest structural associations such as the left caudal anterior cingulate cortex (*r* = −0.22, FDR-*P*_spin_ = 0.048).

At the subcortical level, the combined SUD group showed robust functional epicenters in dorsal striatal and limbic nuclei (Supplementary **Table 15**), including the left putamen (*r* = −0.53, FDR-*P*_shuff_ = 0.001), left caudate (*r* = −0.52, FDR-*P*_shuff_ = 0.001), left thalamus (*r* = −0.51, FDR-*P*_shuff_ = 0.002), left pallidum (*r* = −0.47, FDR-*P*_shuff_ = 0.003), left amygdala (r = −0.47, FDR-*P*_shuff_ = 0.003), and left hippocampus (*r* = −0.46, FDR-*P*_shuff_ < 0.0001). Structural subcortical epicenters were weaker, with the strongest associations observed in the left pallidum (*r* = −0.36, FDR-*P*_shuff_ = 0.004), right thalamus (*r* = −0.20, FDR-*P*_shuff_ = 0.023), and right putamen (*r* = −0.08, FDR-*P*_shuff_ = 0.030). Substance-specific analyses recapitulated the combined SUD findings and revealed reproducible functional subcortico-cortical epicenters in alcohol, cocaine, amphetamines, and cannabis, with more modest effects for nicotine. In contrast, opioid use disorder yielded only two significant structural subcortical epicenters, namely the left caudate and right putamen (Supplementary **Table 15**).

#### Fig. 3. SUD-specific cortical and subcortical epicenters

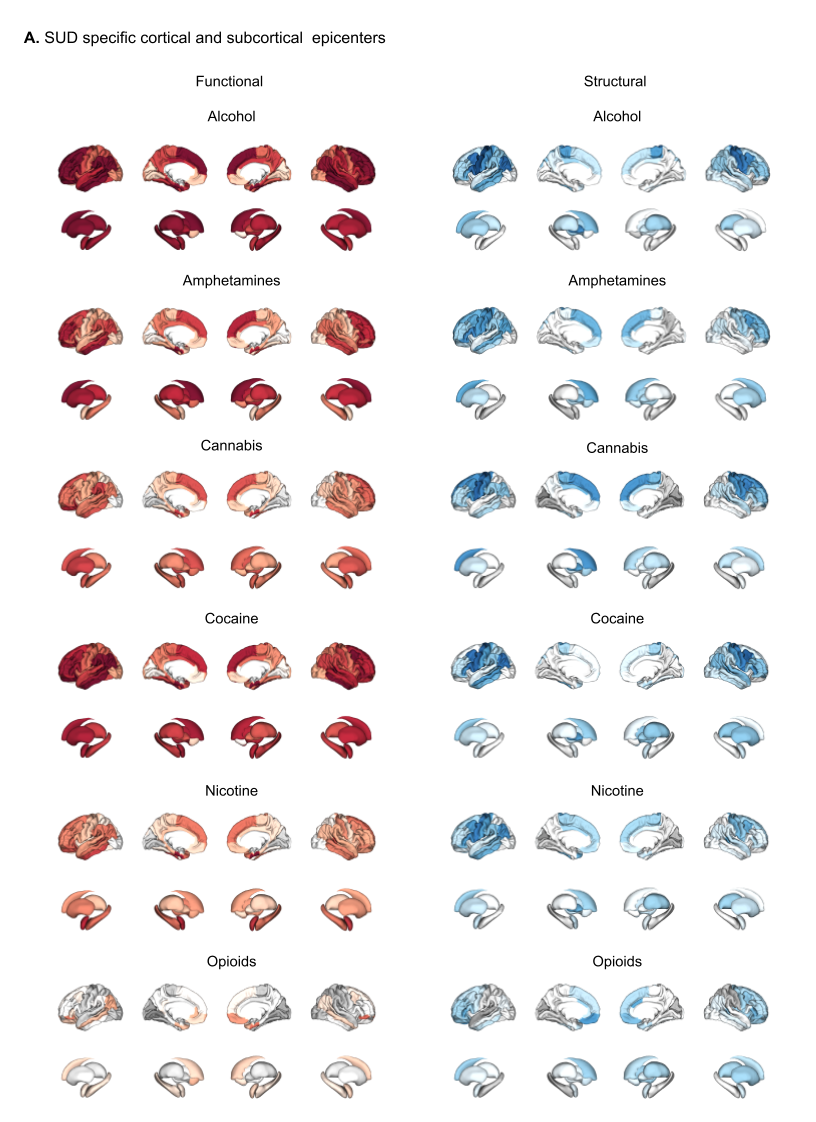

**Supplementary Fig. 3.** (**A**) Functional (red) and structural (blue) cortical and subcortical epicenter maps for each substance use disorder. Epicenters were defined as regions whose normative connectivity profiles showed significant spatial correlation with substance-specific morphometric alteration maps. Functional epicenters primarily involve limbic–temporal and orbitofrontal regions, whereas structural epicenters are more spatially restricted and shift toward frontoparietal and sensorimotor areas.

Abbreviation: SUD = substance use disorder

**Alt text:** Brain maps showing substance-specific functional and structural cortical and subcortical epicenters, highlighting limbic-temporal, orbitofrontal, frontoparietal, and sensorimotor epicenter patterns across substance use disorder categories.

#### Cross-correlation analyses and overlap of epicenters in SUD

To assess convergence across substances, we compared cortical and subcortical alteration maps of each SUD with the combined SUD profile (Supplementary **Table 16**). Pairwise cross-correlation matrices and principal component analysis revealed a dominant shared morphometric pattern across SUD categories (Supplementary **Fig. 2**). Cortical functional maps showed strong similarity for alcohol, amphetamines, cannabis, cocaine, and nicotine (*r* = 0.82–0.98; all FDR-*P*_spin_ < 0.001), whereas opioids exhibited only moderate correspondence (*r* = 0.43–0.72; FDR-*P*_spin_ = 0.001–0.007). Cortical structural maps revealed a similar pattern, with high cross-correlations for alcohol, amphetamines, cannabis, cocaine, and nicotine (*r* = 0.78–0.97; all FDR-*P*_spin_ < 0.01) and weaker associations for opioids (r ≤ 0.54; FDR-*P*_spin_ ≥ 0.10).

At the subcortical level, functional maps showed very strong correspondence for alcohol and cocaine (*r* = 0.96–0.99; FDR-*P*_spin_ < 0.001), moderate alignment for cannabis (*r* ≈ 0.54; FDR-*P*_spin_ = 0.019), and weaker or non-significant associations for nicotine, amphetamines, and opioids (*r* = −0.38–0.39; FDR-*P*_spin_ ≥ 0.06). Structural maps revealed a similar pattern, with robust cross-correlations among alcohol, cannabis, cocaine, and nicotine (*r* = 0.71–0.998; all FDR-*P*_spin_ < 0.001), and weaker associations for amphetamines and opioids (*r* = 0.32–0.48; FDR-*P*_spin_ = 0.005–0.065) (Supplementary **Table 17**). Together, these findings indicate substantial overlap in the spatial organization of cortical and subcortical morphometric alterations across most substance use disorder categories, with opioids exhibiting comparatively lower cross-correlation strength relative to other substances (Supplementary **Fig. 2**).

#### Unique and overlapping epicenters of SUD

The most consistent overlap of cortical epicenters was observed in limbic–temporal territories, including entorhinal cortex, lateral orbitofrontal cortex, inferior parietal lobule, and inferior temporal cortex, shared across alcohol, amphetamines, cocaine, and cannabis, with more limited and region-specific convergence for nicotine (**Fig. 4C** in the main manuscript). Additional regions, such as banks of superior temporal sulcus and anterior cingulate, also showed cross-substance overlap. Structural cortical epicenters showed less convergence, mainly restricted to alcohol and cocaine, for instance in the precentral gyrus and postcentral gyrus (Supplementary **Table 18)**. Subcortically, functional epicenters across alcohol, cannabis, and cocaine localized to amygdala, hippocampus, caudate, putamen, pallidum, and thalamus, with nicotine showing partial overlap, while the nucleus accumbens emerged as a functional epicenter selectively in cannabis.

Structural subcortical overlap was more restricted, primarily involving the pallidum and caudate for alcohol and cocaine (Supplementary **Table 19**).

### Robustness analyses

#### Age and sex modulation of cortical thickness and subcortical volume in SUD

Age-by-diagnosis and sex-by-diagnosis interaction models were fitted in the combined SUD phenotype to identify cortical and subcortical regions where demographic moderator effects interacted with SUD-related morphometric differences. Age-by-diagnosis interactions on cortical thickness and subcortical volume were evaluated using linear models with false discovery rate correction (Supplementary **Tables 20–21**). Significant interaction effects were observed across several cortical regions, with the strongest effects localizing to frontal, temporal, parietal, and insular cortices. Notable examples included the left superior frontal gyrus (*T* = –5.85, FDR-corrected *P* < 0.0001), left middle temporal gyrus (*T* = –5.68, FDR-corrected *P* < 0.0001), right middle temporal gyrus (*T* = –5.54, FDR-corrected *P* < 0.0001), and right superior frontal gyrus (*T* = –5.42, FDR-corrected *P* < 0.0001), indicating stronger SUD-related cortical reductions at older ages. By contrast, no age-by-diagnosis interaction survived FDR correction in any subcortical region (Supplementary **Table 21**). The cortical SUD-by-age interaction map was further evaluated using hub and epicenter mapping, showing significant associations with cortical functional and structural hubness and with canonical SUD epicenter maps (Supplementary **Fig. 4**; Supplementary **Table 22**).

Sex-by-diagnosis interactions revealed no significant cortical effects after FDR correction across any region (Supplementary **Table 23**). At the subcortical level, significant interactions emerged in the bilateral thalamus (left: *T* = –3.40, FDR-corrected *P* = 0.005; right: *T* = –3.25, FDR-corrected *P* = 0.006) and bilateral putamen (left: *T* = –3.44, FDR-corrected *P* = 0.005; right: *T* = –3.09, FDR-corrected *P* = 0.007), indicating stronger SUD-related volume reductions in males compared with females in these regions (Supplementary Fig. **4**; Supplementary **Table 24**).

Accordingly, hub and epicenter analyses were not further applied to sex effects.

Together, these findings indicate that age-related cortical decline in SUD follows hub vulnerability and epicenter patterns similar to those observed for cross-sectional morphometric differences.

##### Fig. 4. Hub and epicenter mapping of SUD-by-age interaction

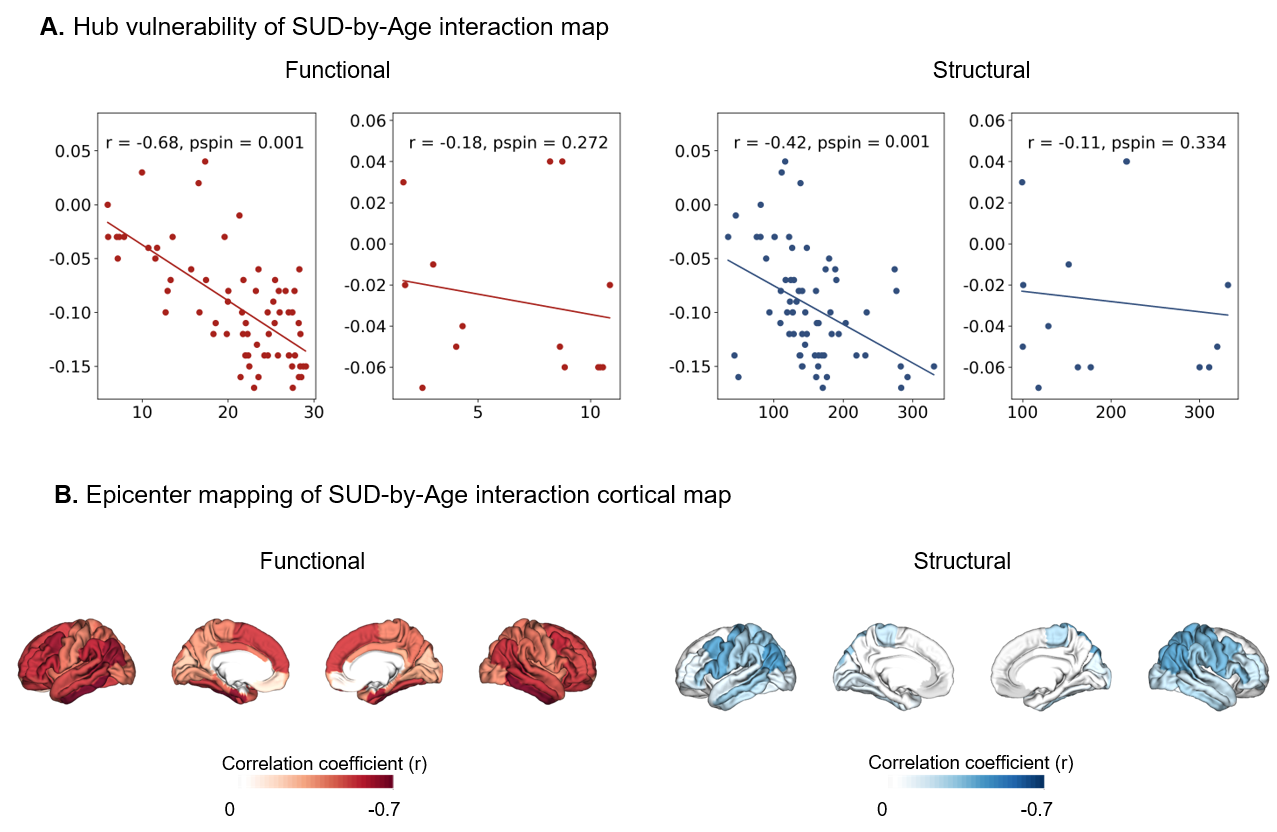

**Supplementary Fig. 4.** (**A**) SUD-by-age–related effects on brain morphology were significantly associated with functional cortical hubness (*r* = –0.68, *P*_spin_ = 0.001) and structural cortical hubness (*r* = –0.42, *P*_spin_ = 0.001), whereas no significant associations were observed for subcortical regions. (**B**) Epicenter mapping of the SUD-by-age cortical interaction effects revealed that the resulting epicenter maps were strongly correlated with the canonical SUD functional (*r* = –0.84, *P*_spin_ < 0.0001) and structural (*r* = 0.62, *P*_spin_ = 0.0035) epicenter maps.

Abbreviations: *P*spin = spin-permutation P-value; *r* = Pearson correlation coefficient; SUD = substance use disorder.

**Alt text:** Brain maps and correlation plots showing that age-related morphometric effects in substance use disorders follow cortical hub vulnerability and epicenter patterns similar to the main substance use disorder morphometric maps.

#### Subject-level cortical differences modelling in SUD

Having established our network-based susceptibility models at the group level, we next examined the extent to which these patterns were expressed at the level of individual participants. For each participant with SUD, cortical thickness values were residualized for age and sex and z-scored relative to the healthy control distribution to generate subject-specific cortical abnormality maps. These individualized maps were spatially correlated with normative functional and structural degree centrality maps to derive hub vulnerability scores. Consistent with the group-level findings, individual patients with SUD showed evidence of hub vulnerability and epicenter organization, albeit with greater inter-individual variability and reduced sensitivity at the single-subject level (Supplementary **Fig. 5**; Supplementary **Table 25**).

To identify individual-level epicenters, each cortical region was tested for whether its normative connectivity profile significantly aligned with the participant’s cortical abnormality map using spin-based spatial permutation testing (*P*_spin_ < 0.05). Aggregated epicenter maps were generated by calculating, for each region, the proportion of individuals in whom that region was identified as an epicenter. Group differences in hub vulnerability and epicenter frequency were evaluated using chi-square tests (Supplementary **Table 25**).

Despite this heterogeneity, even at the individual level, patients with SUD exhibited a significantly higher likelihood of hub vulnerability.

Functional hub vulnerability was observed in 24.2% of SUD patients versus 18.5% of controls (chi-squared statistic [χ²] = 19.1, degrees of freedom [df] = 1, *p* < 0.001), while structural epicenters were identified in 22.4% versus 14.9%, respectively (χ² = 41.5, df = 1, *P* < 0.001). Moreover, the group-level epicenter pattern could be replicated at the individual level above chance levels; epicenter likelihood, as quantified by the regional χ² statistics derived from subject-level comparisons (Supplementary **Table 25**; Supplementary **Fig. 5**) was significantly correlated with functional (*r* = –0.72, 95% CI [–0.82, –0.58], *P*_spin_ < 0.001) and structural (*r* = –0.59, 95% CI [–0.73, –0.41], *P*_spin_ < 0.001) group-level analyses.

Together, these findings support a convergence between group-level network vulnerability patterns and their expression across individuals with SUD, while preserving substantial inter-individual variability. As illustrated in Supplementary **Fig. 5**, these findings are reflected in two complementary analyses: (i) distributions of patient-level correlations between cortical differences maps and cortical hubness (degree centrality), showing a higher proportion of significant associations in SUD compared to controls, and (ii) cortical maps indicating the percentage of individuals for whom each region qualified as a functional or structural epicenter.

##### Fig. 5. Individual-level network modelling analysis in SUD

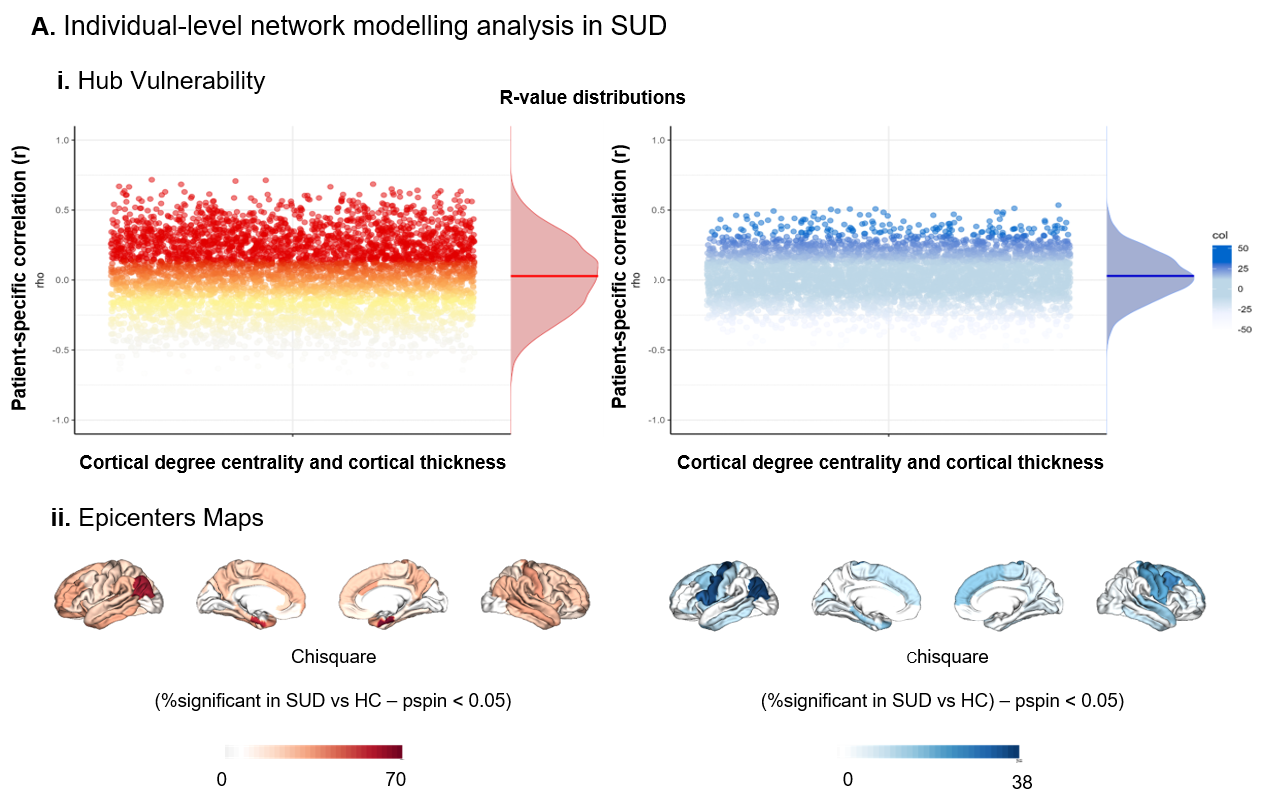

**Supplementary Fig. 5**. (**A**) *i*. Distribution of individual functional (left) and structural (right) hub vulnerability estimates in patients with SUD; (**A**) *ii*. Brain projection of regional χ²-values of epicenter likelihood in individuals with SUD vs healthy controls. The percentage of individuals with SUD and healthy controls for whom each region qualifies as a significant functional (left) or structural (right) epicenter was compared with the χ² test.

Abbreviations: HC = healthy controls; SUD = substance use disorder; vs = versus; χ² = chi-squared statistic.

**Alt text:** Distribution plots and cortical maps showing individual-level hub vulnerability and epicenter likelihood in patients with substance use disorders compared with healthy controls.

#### Robustness of SUD-specific morphometric differences to age distribution

Because of the strong association between age and brain morphometry, as well as the diagnosis-by-age interaction observed in SUD, we tested whether the comparatively larger case–control effects in alcohol and cocaine use disorders could be explained by their older age profiles.

We repeated the case–control analysis under an age harmonization weighting designed to downshift alcohol/cocaine toward a younger reference while avoiding age inflation in the other SUD subgroups. Within the filtered non-comorbid subset (age ≤ preset maximum; complete age, sex, and intracranial volume (ICV) data), we defined an anchor reference from amphetamines/cannabis/nicotine/opioids and computed its mean age. For each subgroup, we resampled 60% of available cases without replacement across bootstrap iterations. Amphetamines/cannabis/nicotine/opioids were sampled uniformly (preserving their natural age distributions). For alcohol/cocaine only, we used weighted rejection sampling to preferentially select younger participants and accepted draws only when the achieved mean age matched a lower target mean age within a ±2-year tolerance; targets were defined one-sided (set to the anchor mean, but never allowed to exceed the subgroup’s natural mean) to ensure alcohol/cocaine could only shift younger. For each accepted case draw, controls were sampled without replacement to match the case sample’s age-bin counts (age-aligned case–control sampling). Regional effects were re-estimated using linear models adjusted for age, sex, and ICV, and summarized per iteration by the median Cohen’s *d* across regions. Under one-sided age harmonization, alcohol and cocaine retained the largest cortical effects despite younger target mean ages (target mean ≈ 30.47 years; median d: alcohol ≈ −0.19, cocaine ≈ −0.20), exceeding amphetamines (≈ −0.09) and comparable to or larger than cannabis/nicotine/opioids (≈ −0.14 to −0.15; Supplementary **Table 26**; Supplementary **Fig. 6**). Subcortically, alcohol remained among the largest effects (≈ −0.13), with cocaine/cannabis/opioids similar (≈ −0.11 to −0.12) and amphetamines/nicotine smaller (≈ −0.08 to −0.09; Supplementary **Table 27**; Supplementary **Fig. 6**). Relative to the original full-cohort estimates (median d; cortex: −0.25, −0.11, −0.18, −0.20, −0.17, −0.15; subcortex: −0.20, −0.08, −0.09, −0.06, −0.11, −0.11 for alcohol/amphetamines/cannabis/cocaine/nicotine/opioids), the age-downshifted bootstrap largely preserved the qualitative ranking of subgroup effects.

These results indicate that higher alcohol- and cocaine-related effect magnitudes were not explained solely by age distribution or SUD-by-age interaction.

##### Fig. 6. Stability of median morphometric effect sizes under age harmonization

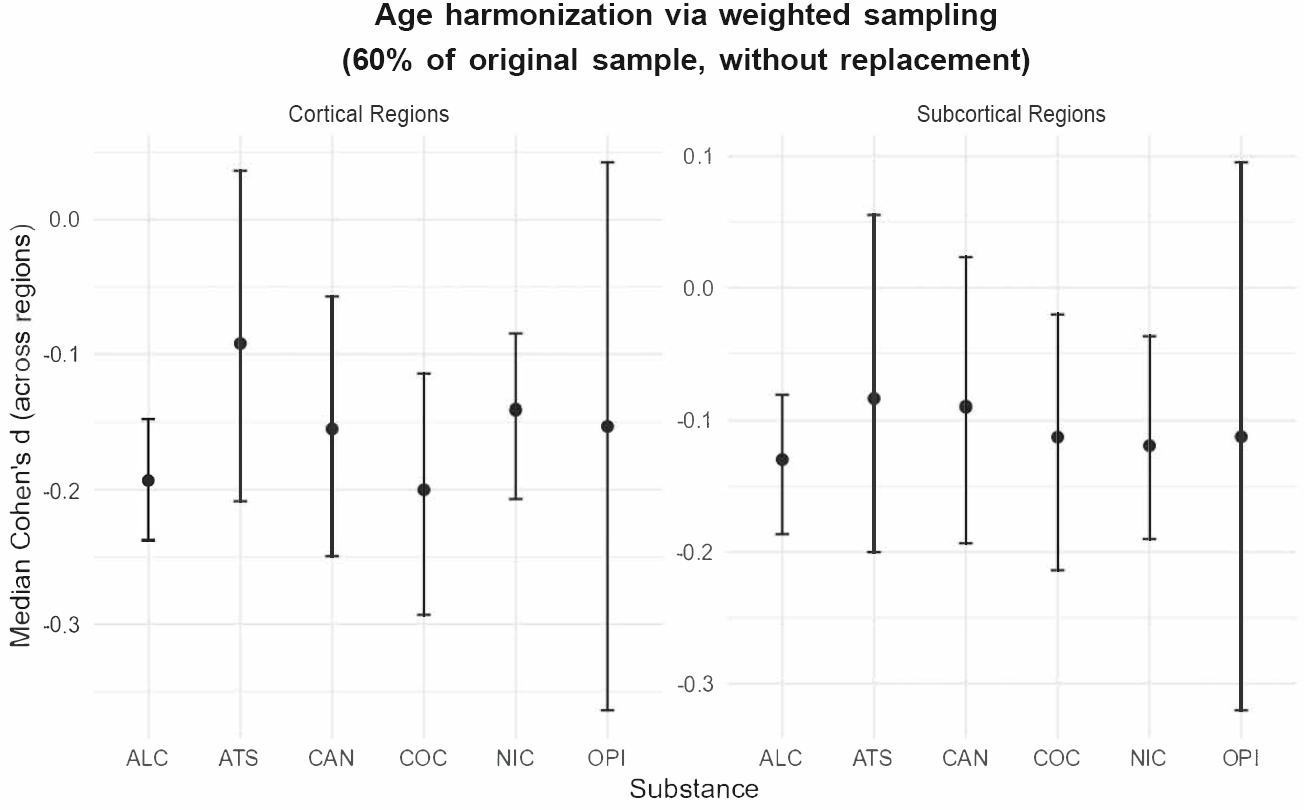

**Supplementary Fig. 6.** Median Cohen’s *d* values across cortical (left) and subcortical (right) regions are shown for each substance use disorder under age harmonization. For each subgroup, 60% of available non-comorbid cases were resampled without replacement across bootstrap iterations; alcohol and cocaine were downshifted toward a lower target mean age via weighted rejection sampling, whereas amphetamines, cannabis, nicotine, and opioids were sampled uniformly. Points indicate the median effect size across regions, and error bars denote 95% bootstrap confidence intervals; values are shown separately for cortical and subcortical metrics. Under age-downshifted resampling, alcohol and cocaine retain among the largest median effect magnitudes, indicating that subgroup differences are not explained solely by older age distributions.

Abbreviations: ALC = alcohol use disorder; ATS = amphetamine-type stimulant use disorder; CAN = cannabis use disorder; CI = confidence interval; COC = cocaine use disorder; NIC = nicotine use disorder; OPI = opioid use disorder; SUD = substance use disorder. Cohen’s d = standardized mean difference.

**Alt text:** Plots showing median cortical and subcortical morphometric effect sizes across substance use disorder groups after age harmonization, demonstrating that alcohol and cocaine retain among the largest effects after age downshifting.

#### Robustness of SUD-specific morphometric differences to sample size

To evaluate whether the observed differences in case–control effect sizes across substance-use subgroups were driven by unequal sample sizes rather than true underlying neuroanatomical variation, we performed a bootstrap analysis with equalized sample sizes across a range of matched *n* values. For each SUD group, cases and matched controls were repeatedly resampled with replacement to fixed target sample sizes ranging from n = 25 to n = 300, with 500 bootstrap iterations per setting. Substances with smaller original cohort sizes, including opioids and amphetamines, were upsampled with replacement so that all groups could be evaluated on the same controlled sample-size grid. We found the magnitude and relative pattern of subgroup effect sizes for alcohol, amphetamines, cannabis, cocaine, nicotine and opioids were highly stable across all resampled sample sizes, with median effects differing only minimally between the original full cohort (median Cohen’s *d* across regions; cortex: –0.25, –0.11, –0.18, –0.20, –0.17, –0.15; subcortex: –0.20, –0.08, –0.09, –0.06, –0.11, –0.11) and the bootstrapped equal-N analyses (median Cohen’s *d* across regions; cortex: –0.23, –0.09, –0.16, –0.20, –0.14, –0.16; subcortex: –0.14, –0.08, –0.09, –0.08, –0.12, –0.10; averaged across matched sample sizes ranging from n = 25 to n = 300).

The central tendency of effect-size estimates across *n* remained stable for both cortical and subcortical metrics, with narrower confidence intervals at higher sample sizes (Supplementary **Fig. 7**). Thus, the relative pattern and stability of effect magnitudes across SUD subgroups cannot be attributed to sample size differences.

##### Fig. 7. Stability of median effect sizes across sample sizes

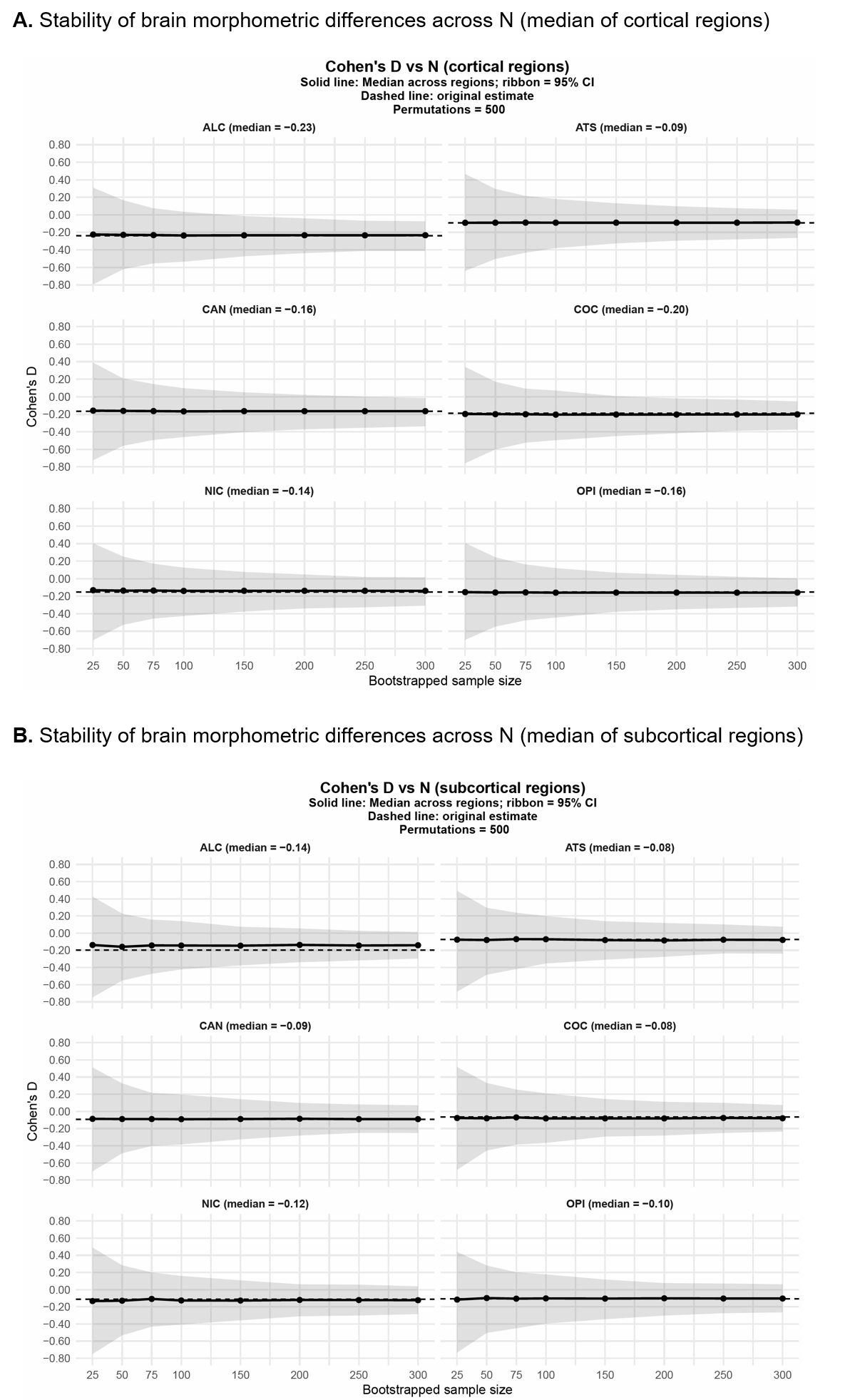

**Supplementary Fig. 7.** Median Cohen’s *d* values across cortical (**A**) and subcortical (**B**) regions are shown for each substance use disorder across bootstrapped equal-*N* subsamples (*n* = 25–300 per group). Solid lines indicate the median effect size across regions, shaded ribbons represent 95% bootstrap confidence intervals, and dashed lines denote the original full-sample estimates. Across the full range of subsampled sizes, median effect sizes remain highly stable in magnitude and direction, with increasing precision at larger *N*, indicating that observed group differences are robust to variation in sample size and not driven by unequal group sizes.

Abbreviations: ALC = alcohol use disorder; ATS = amphetamine-type stimulant use disorder; CAN = cannabis use disorder; CI = confidence interval; COC = cocaine use disorder; *N* = sample size; NIC = nicotine use disorder; OPI = opioid use disorder; SUD = substance use disorder. Cohen’s *d* = standardized mean difference.

**Alt text:** Line plots showing the stability of median cortical and subcortical effect sizes across bootstrapped equal sample sizes for each substance use disorder group.

#### Robustness of SUD-specific morphometric difference patterns to sample size

To complement the median effect-size analysis, we next evaluated the stability of the covariance structure of morphometric differences as a function of sample size. For each substance, we generated bootstrap case–control maps at increasing *N* (25–300) and computed, per iteration, the spatial correlation between the region-wise Cohen’s *d* vector at a given n and the reference pattern derived from the full-sample estimate (*N* = 300). This procedure was repeated for 500 bootstrap iterations at each sample size, and spatial stability was summarized using the median Pearson correlation, 95% bootstrap confidence interval, and standard deviation across iterations. Across substances, median correlations increased monotonically with sample size, showing lower and more variable correspondence at small *N* and progressively stabilizing as *N* approached 300 (Supplementary **Fig. 8**). As expected, the covariance structure was more sensitive to sample size than the median Cohen’s d values, because it depends on the full regional ordering rather than a single summary statistic averaged across regions. Nonetheless, all substances showed convergence toward the reference pattern with increasing *N* (e.g., *n* = 75 → all *r* ≥ 0.5; *n* = 150 → all *r* ≥ 0.6), and confidence interval widths narrowed accordingly. These results indicate that while regional effect-size ordering is less stable at small sample sizes, the underlying spatial correlation structure becomes reproducible at moderate to large *N*, supporting the robustness of the observed morphometric patterns.

##### Fig. 8. Stability of covariance structure patterns across sample sizes

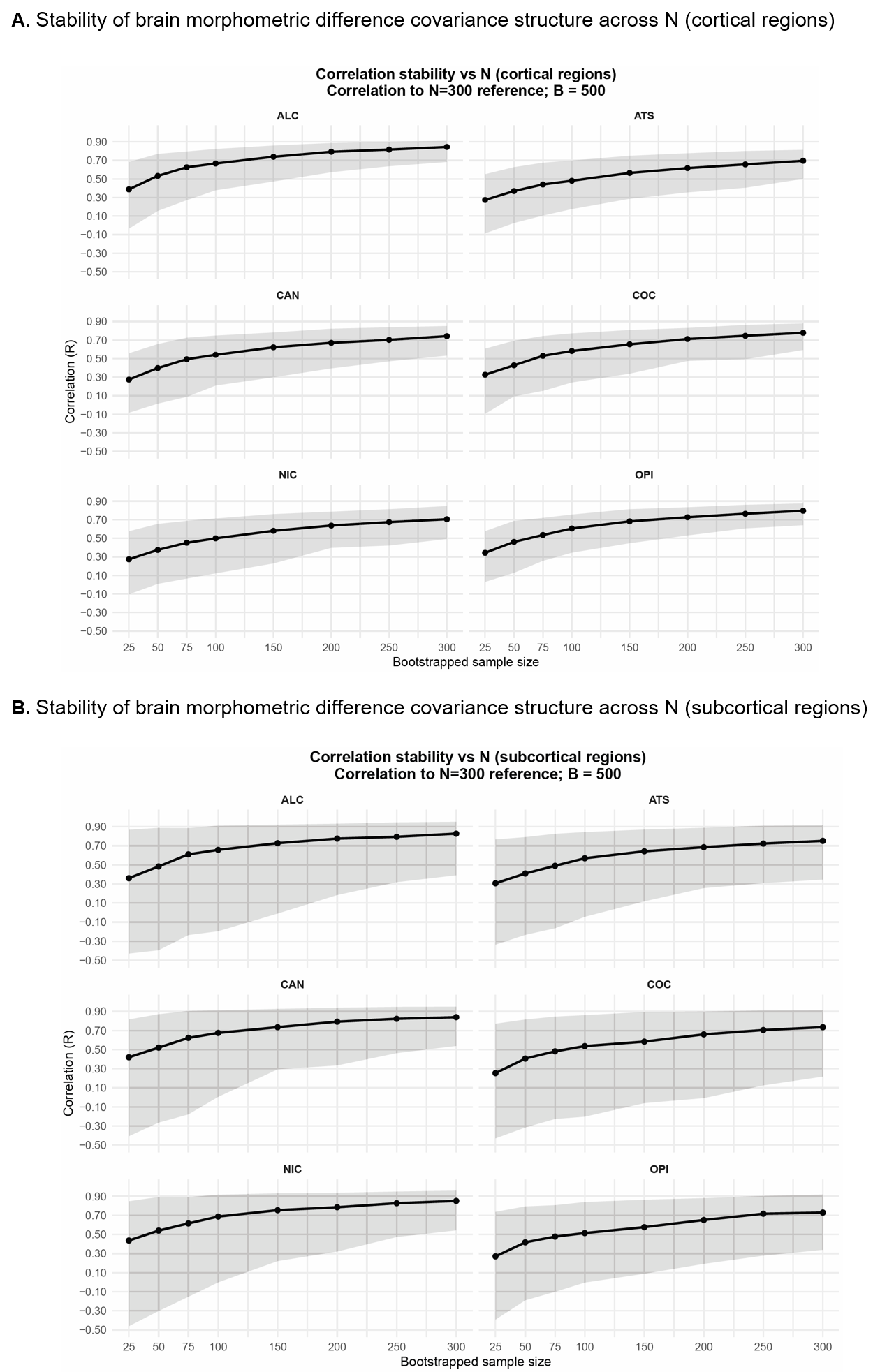

**Supplementary Fig. 8.** Correlation coefficients between bootstrapped regional Cohen’s *d* vectors and a fixed large-*N* reference map (*n* = 300) across sample sizes (*n* = 25–300). Curves show median Pearson correlations (r) with 95% bootstrap confidence intervals. Spatial correspondence increases monotonically with sample size, with higher variability at small *N* and narrower confidence intervals at larger N. From moderate sample sizes, correlation values stabilize (e.g., *n* = 75 → all *r* ≥ 0.5; n = 150 → all *r* ≥ 0.6) with the ground truth, indicating that the spatial pattern of regional effects is robust to sample size differences.

Abbreviations: ALC = alcohol use disorder; ATS = amphetamine-type stimulant use disorder; CAN = cannabis use disorder; CI = confidence interval; COC = cocaine use disorder; N = sample size; NIC = nicotine use disorder; OPI = opioid use disorder; r = Pearson correlation coefficient; SUD = substance use disorder. Cohen’s d = standardized mean difference.

**Alt text:** Line plots showing increasing spatial stability of regional morphometric effect-size patterns across bootstrapped sample sizes, with stronger correspondence to the reference map at larger sample sizes.

**Robustness of SUD-specific brain morphometric differences to control group sample**

Lastly, to assess whether substance-specific morphometric differences depended on the composition of the healthy control group, we repeated the case–control analyses using the study-specific control samples provided in the original ENIGMA SUD datasets. Study-specific healthy control samples included alcohol: *n* = 765, amphetamines: *n* = 233, cannabis: *n* = 247, cocaine: *n* = 291, nicotine: *n* = 327, and opioids: *n* = 88. In the primary analysis, individuals with a single diagnosed SUD were compared to the full pooled healthy control cohort (*N* = 1,951). This primary analysis corresponded to the single-SUD groups shown in **Fig. 2A** in the main manuscript and Supplementary **Table 2** (alcohol: *n* = 902; amphetamines: *n* = 178; cannabis: *n* = 286; cocaine: *n* = 278; nicotine: *n* = 600; opioids: *n* = 68). Region-wise cortical Cohen’s *d* values derived from this pooled control approach are reported in Supplementary **Table 28**.

First, we compared substance-specific control groups to individuals with one or more comorbid SUD diagnoses (Split_HC_comorbid; Supplementary **Table 29**), representing the most inclusive case definition. The comorbid SUD samples were defined according to Supplementary **Table 2** and included alcohol: *n* = 1,216; amphetamines: *n* = 185; cannabis: *n* = 288; cocaine: *n* = 404; nicotine: *n* = 662; and opioids: *n* = 92. Second, we compared the same substance-specific control groups to individuals with a single SUD diagnosis (Split_HC_noncomorbid; Supplementary **Table 30**), thereby matching the patient definition used in the primary pooled analysis.

For all analyses, region-wise Cohen’s *d* values were recalculated using the same covariate-adjusted linear models as in the main analysis, and spatial correspondence between maps was evaluated using Pearson correlations across regions. Correlations show high spatial overlap across approaches (range = 0.67–0.99), with slightly reduced similarity for substances with smaller control samples, while preserving the overall spatial pattern of cortical alterations across methods (Supplementary **Fig. 9**). This approach allowed us to determine how differences in control-sample origin and sample size influence the stability of substance-specific morphometric patterns.

##### Fig. 9. Correlation matrix across SUD effect-size maps under three control-group methods

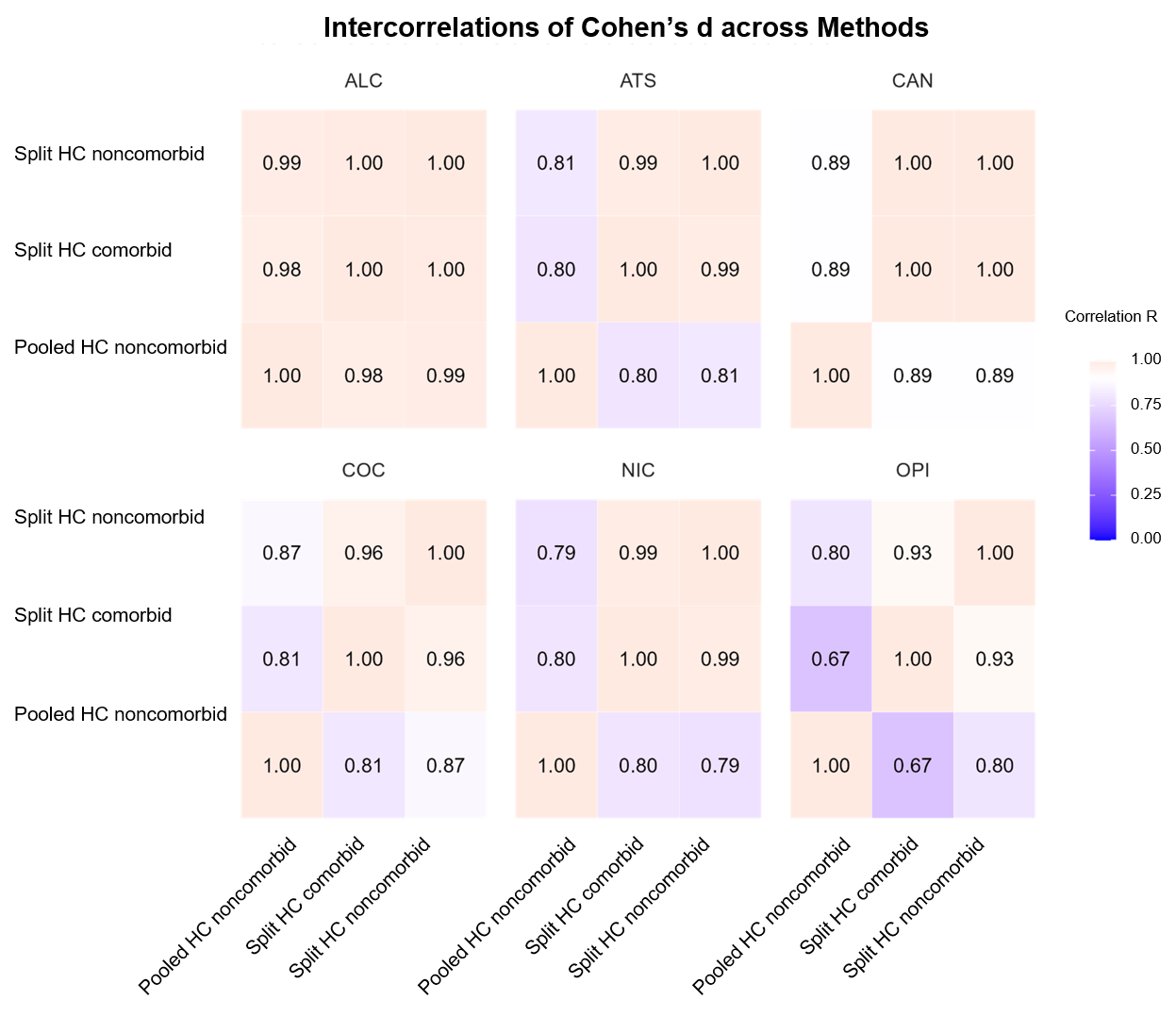

**Supplementary Fig. 9.** Pairwise Pearson correlations between cortical Cohen’s *d* maps obtained from three analytical approaches. Method 1 (Pooled_HC_noncomorbid) corresponds to the primary analysis comparing individuals with a single SUD diagnosis to the full pooled healthy control sample (*N* = 1,951; see Supplementary **Table 28**). Method 2 (Split_HC_noncomorbid) compares individuals with a single SUD diagnosis to study-specific healthy control samples (see Supplementary **Table 30**). Method 3 (Split_HC_comorbid) compares individuals with one or more comorbid SUD diagnoses to the same study-specific healthy control samples (see Supplementary **Table 29**). Warmer colours indicate stronger spatial agreement between effect-size patterns. Overall, correlations indicate high spatial concordance across approaches (range = 0.67–0.99), with slightly reduced similarity for substances with smaller control samples.

Abbreviations: ALC = alcohol use disorder; ATS = amphetamine-type stimulant use disorder; CAN = cannabis use disorder; COC = cocaine use disorder; HC = healthy controls; N = sample size; NIC = nicotine use disorder; OPI = opioid use disorder; r = Pearson correlation coefficient; SUD = substance use disorder. Cohen’s d = standardized mean difference. Method labels: Pooled_HC_noncomorbid = pooled healthy control sample with non-comorbid SUD cases; Split_HC_noncomorbid = study-specific healthy control samples with non-comorbid SUD cases; Split_HC_comorbid = study-specific healthy control samples with comorbid SUD cases.

**Alt text:** Correlation matrix showing high spatial agreement between cortical effect-size maps obtained using pooled healthy controls, study-specific non-comorbid controls, and study-specific comorbid control-group approaches.

**Disease epicenter mapping of six psychiatric disorders**

We identified cortical and subcortical epicenters across six psychiatric disorders: schizophrenia (SCZ), bipolar disorder (BD), major depressive disorder (MDD), obsessive–compulsive disorder (OCD), autism spectrum disorder (ASD), and attention-deficit/hyperactivity disorder (ADHD), using the same epicenter mapping approach as in the SUD analyses (see Methods; Supplementary **Tables 31–34**).

In SCZ, the epicenter pattern was dominated by a temporo-limbic axis, with the most representative functional epicenters located in the banks of the superior temporal sulcus and the entorhinal cortex bilaterally (e.g., left banks of the superior temporal sulcus: r = −0.55, *P*_spin_ < 0.0001; left entorhinal cortex: *r* = −0.63, *P*_spin_ < 0.0001; Supplementary **Table 31**). These regions consistently anchored the spatial distribution of cortical thickness alterations and were complemented by inferior temporal involvement. Structurally, convergent epicenters extended beyond temporal cortex into parieto-frontal regions, notably the inferior parietal cortex and inferior frontal gyrus, indicating a broader fronto-temporal propagation of morphometric vulnerability (Supplementary **Table 32**).

At the subcortical level, SCZ showed a robust striato-limbic signature, with bilateral involvement of the amygdala, caudate nucleus, hippocampus, and putamen as functional epicenters, whereas structural subcortical epicenters were more limited (Supplementary **Tables 33–34**).

In BD, cortical epicenters were characterized by a predominantly prefrontal and fronto-striatal organization, rather than a focal temporo-limbic pattern. The most representative functional epicenters were observed in the caudal middle frontal gyrus and orbitofrontal cortex (e.g., left caudal middle frontal gyrus: *r* = −0.57, *P*_spin_ < 0.0001; left lateral orbitofrontal cortex: *r* = −0.58, *P*_spin_ < 0.0001; Supplementary **Table 31**), reflecting a diffuse prefrontal anchoring of cortical alterations. Structurally, this pattern converged on dorsolateral prefrontal regions, with the rostral middle frontal gyrus emerging as a key epicenter (Supplementary **Table 32**).

In MDD, the epicenter organization was more circumscribed and less spatially extensive than in schizophrenia or bipolar disorder. Cortically, the most informative structural epicenters were confined to the caudal anterior cingulate cortex bilaterally (e.g., left caudal anterior cingulate: *r* = −0.37, *P*_spin_ = 0.008; Supplementary **Table 32**), highlighting a focal midline vulnerability rather than a distributed cortical network. In contrast, the subcortical compartment played a more prominent role, with the nucleus accumbens emerging as the key epicenter, both functionally and structurally (functional: left accumbens *r* = −0.33, *P*_spin_ = 0.009; structural: bilateral accumbens *r* ≤ −0.41, *P*_spin_ ≤ 0.001; Supplementary **Tables 33–34**). This pattern emphasizes a ventral striatal–limbic anchoring of morphometric alterations in depression.

In OCD, the epicenter architecture was organized around a fronto-striatal control network, with a relative sparing of temporo-limbic regions compared to schizophrenia. The most representative functional cortical epicenters were located in the caudal middle frontal gyrus bilaterally (e.g., left caudal middle frontal gyrus: *r* = −0.49, *P*_spin_ < 0.0001; Supplementary **Table 31**), highlighting a dominant dorsolateral prefrontal anchoring of cortical alterations. Structurally, convergence was observed in the rostral middle frontal gyrus, consistent with involvement of executive control regions (Supplementary **Table 32**).

At the subcortical level, OCD showed a selective dorsal striatal signature, with the caudate nucleus acting as the principal functional epicenter (e.g., right caudate: *r* = −0.44, *P*_spin_ = 0.001; Supplementary **Table 33**), accompanied by additional involvement of the thalamus. Structural subcortical epicenters extended to the nucleus accumbens and putamen, reinforcing the centrality of fronto-striato-thalamic circuits in OCD-related morphometric vulnerability (Supplementary **Table 34**).

In ASD, epicenter patterns were sparser and less spatially coherent across both cortical and subcortical compartments. Functionally, the most informative cortical epicenters were located in posterior and sensory–association regions, particularly the cuneus (e.g., left cuneus: *r* = 0.49, *P*_spin_ = 0.008; Supplementary **Table 31**), indicating a shift away from canonical fronto-limbic anchoring. Structural cortical epicenters were limited and heterogeneous, without a dominant prefrontal or temporal focus (Supplementary **Table 32**).

At the subcortical level, ASD showed weak and inconsistent epicenter involvement, with no single nucleus emerging as a robust cross-modal hub, underscoring the relative decentralization of morphometric alterations in this disorder (Supplementary **Tables 33–34**).

ADHD exhibited the least pronounced epicenter organization among the psychiatric disorders examined. Cortically, functional epicenters were modest and primarily localized to posterior cortical regions, including the cuneus and lateral occipital cortex (e.g., left cuneus: *r* = 0.41, *P*_spin_ = 0.009; Supplementary **Table 31**), rather than fronto-striatal control areas. Structural cortical epicenters were rare and did not converge on a consistent anatomical substrate (Supplementary **Table 32**).

Subcortically, ADHD showed no robust epicenters across modalities, with only weak, non-convergent associations observed across nuclei, consistent with a diffuse and low-amplitude pattern of morphometric alterations (Supplementary **Tables 33–34**). Transdiagnostic cortical and subcortical epicenter correlations between SUD and psychiatric disorder maps are reported in Supplementary **Tables 35–36**.

### Overlap of epicenters between SUD and six psychiatric disorders

Overlap analyses identified the regional convergence of epicenters between substance use disorders (SUD) and psychiatric disorders, based on the spatial overlap of FDR-*P*_spin_ significant epicenters in each disorder (Supplementary **Tables 37**–**38**).

At the cortical level, functional epicenters in SUD overlapped predominantly with those observed in SCZ and BD, involving bilateral temporal and paralimbic regions, including the banks of the superior temporal sulcus, entorhinal cortex, inferior parietal cortex, middle and inferior temporal gyri, as well as distributed prefrontal regions such as the caudal and rostral middle frontal gyrus, pars opercularis, pars orbitalis, pars triangularis of the inferior frontal gyrus, lateral orbitofrontal cortex, and the frontal and temporal poles. Structural cortical overlap was more limited and was primarily observed with BD, encompassing the bilateral caudal middle frontal gyrus, left inferior parietal cortex, and right pars triangularis of the inferior frontal gyrus.

Overlap with MDD was rare and restricted to functional epicenters in the frontal poles and temporal pole, with no consistent structural cortical overlap.

At the subcortical level, functional epicenters associated with SUD overlapped mainly with SCZ, and to a lesser extent with BD (Supplementary **Table 38**). Shared regions included the bilateral caudate nucleus and left putamen, while SCZ-specific overlap additionally involved the bilateral amygdala, hippocampus, and left pallidum.

No subcortical epicenter overlap was observed with MDD, ASD, or ADHD.

Overall, these overlap patterns delineate a core set of limbic and striatal regions acting as shared epicentral hubs between SUD and SCZ, with more limited convergence with BD, and minimal overlap with mood (MDD) or neurodevelopmental disorders (ASD, ADHD).

### Neurotransmitter receptor-density mapping

The complete cortical receptor-density matrix used for the neurotransmitter analyses is provided in Supplementary **Table 39**, including z-scored densities for 20 neurotransmitter receptor and transporter maps across all 68 DKT cortical regions. These standardized maps were used to quantify whether SUD-related cortical alteration patterns preferentially aligned with specific neurotransmitter systems.
